# Genome-wide association study of susceptibility to acute respiratory distress syndrome

**DOI:** 10.1101/2025.02.11.25322045

**Authors:** Beatriz Guillen-Guio, Eva Suarez-Pajes, Eva Tosco-Herrera, Tamara Hernandez-Beeftink, Jose Miguel Lorenzo-Salazar, Diana Chang, Rafaela González-Montelongo, Luis A. Rubio-Rodríguez, Olivia C. Leavy, Richard J. Allen, Almudena Corrales, Raquel Cruz, Miguel Bardají-Carrillo, Angel Carracedo, Eduardo Tamayo, V. Eric Kerchberger, Lorraine B. Ware, Brian L. Yaspan, Markus Scholz, André Scherag, Jesús Villar, Louise V. Wain, Carlos Flores

**Author notes:** Corresponding author: Dr. Carlos Flores., Research Unit, Hospital Universitario Nuestra Señora de Candelaria, Santa Cruz de Tenerife, Spain. Equal contribution as senior authors (Louise V. Wain, Carlos Flores).

## Abstract

**Introduction:** Acute respiratory distress syndrome (ARDS) is a severe inflammatory process of the lung, often due to sepsis, and poses significant mortality burden in intensive care units. Here we conducted the largest genome-wide association study (GWAS) of sepsis-associated ARDS to identify novel genetic risk loci that can help guide the development of new therapeutic options.

**Methods:** We performed a case-control GWAS in 716 patients with sepsis-associated ARDS and 4,399 at-risk sepsis controls from three independent studies. Results were meta-analysed across the three studies, with significance set at *p*<5×10^-8^. Suggestive associations were declared for variants exhibiting consistent effects, likely to replicate and nominal significance (*p*<0.05) in all three studies. Prioritised loci were subjected to Bayesian fine mapping, *in-silico* functional assessments, and gene-based rare variant collapsing analysis using whole exome sequencing (WES) data. Two independent studies with 430 ARDS cases and 1,398 controls served as replication samples.

**Results:** We identified a variant showing genome-wide significant association with sepsis-associated ARDS risk intergenic to *ANKRD31* and *HMGCR*, previously linked to cholesterol metabolism. Suggestive associations were found for eight other variants. The rare exonic variant analysis showed associations between *HMGCR* and *POC5* and sepsis-associated ARDS at nominal level (*p*<0.05). While no nominal significance was achieved in the two additional validation cohorts, three variants exhibited a consistent direction of effects across all 5 studies.

**Conclusion:** A common variant intergenic to *ANKRD31* and *HMGCR* was associated with sepsis-associated ARDS risk, suggesting a link between cholesterol metabolism and ARDS risk. Validation in independent studies is needed.

## Introduction

Acute respiratory distress syndrome (ARDS) is a severe lung condition with an overall hospital mortality of about 40% (Bellani et al., 2016). It develops due to injury to the capillary alveolar membrane, which can be triggered by direct or indirect causes, including pulmonary and non-pulmonary sepsis, severe pneumonia, major trauma, and blood transfusion, among others (Force et al., 2012). ARDS is characterised by a rapid onset of lung inflammatory damage, resulting in hypoxemia and acute respiratory failure. Furthermore, ARDS survivors frequently manifest long-term complications, including pulmonary fibrosis development. The diagnosis of ARDS still relies on clinical and imaging criteria (Villar et al., 2023).

ARDS is a medical emergency that requires immediate management in intensive care units (ICUs). The treatment of ARDS primarily involves supportive care such as mechanical ventilation to improve oxygenation, prone positioning, and medications to address underlying causes and manage symptoms. There is a lack of specific lung-directed pharmacological treatments with demonstrated benefit for ARDS patients in clinical trials (Shaw et al., 2019). Thus, seeking effective treatments and specific prognostic methods is crucial for enhancing the survival outcomes of ARDS patients.

Leveraging genomic information could hold the key for guiding the search for future ARDS treatments, as it supports clinical trial efficiency by identifying drug targets with genetic support (Nelson et al., 2015). The genetics of ARDS is complex and not yet fully understood. However, there is evidence supporting the central role of host genetic factors in the development and severity of ARDS (Suarez-Pajes et al., 2023). Genome-wide association studies (GWAS) have been used to identify genetic variants contributing to the risk of this syndrome typically by comparing genetic variation between cases and controls. We recently completed a GWAS of sepsis-associated ARDS in a two-stage study comprising 1,935 ICU patients (633 ARDS cases and 1,302 controls with sepsis) (Guillen-Guio et al., 2020). This analysis identified a novel genome-wide significant locus associated with ARDS susceptibility in a regulatory region of the promoter region of the *FLT1* gene, encoding the vascular endothelial growth factor receptor 1.

Here we aimed to detect additional genetic loci involved in sepsis-associated ARDS to improve our understanding of the syndrome. To our knowledge, we have conducted the largest and more comprehensive GWAS of susceptibility to sepsis-associated ARDS by meta-analysing case-control genotype data from three different studies, followed by the integration of exome sequencing data and findings from two additional studies.

## Methods

### Study design

We performed a GWAS meta-analysis on 5,115 critically ill patients, 82.8% with sepsis, comprising 716 ARDS cases and 4,399 at-risk controls from three independent studies (**Figure 1**): the GENetics of SEPsis-induced ARDS Network (GEN-SEP), the Critical Care Trials Group of the German Sepsis Competence Network (SepNet), and UK Biobank (UKBB). We included a total of 805 patients from the GEN-SEP cohort (304 ARDS cases and 501 at-risk controls), 740 patients from SepNet (91 ARDS cases and 649 at-risk controls), and 3,570 patients from UKBB (321 ARDS cases and 3,249 at-risk controls). Of these, 1,330 patients were included in the prior GWAS (590 from GEN-SEP and 740 from SepNet) (Guillen-Guio et al., 2020). The GEN-SEP and SepNet studies comprised cases of clinically defined sepsis-associated ARDS (Berlin definition) (Force et al., 2012) and controls with sepsis who did not develop ARDS. The UKBB study was based on electronic health record data, encompassing 75.4% of patients with ARDS associated with ICD-10 codes indicative of sepsis (n=242) and 24.6% with ARDS associated with other causes (n=79). ICD-10 codes used to determine the ARDS development causes are provided in **Table S1**. UKBB controls were critically ill patients who did not develop ARDS (75.4% with ICD-10 codes indicative of sepsis). All individuals from the GEN-SEP and SepNet cohorts were of European ancestry, while more than 92% of individuals in UKBB were of European ancestry (**Table S2**). Further details of the three studies are provided in the appendix.

**Figure 1.**
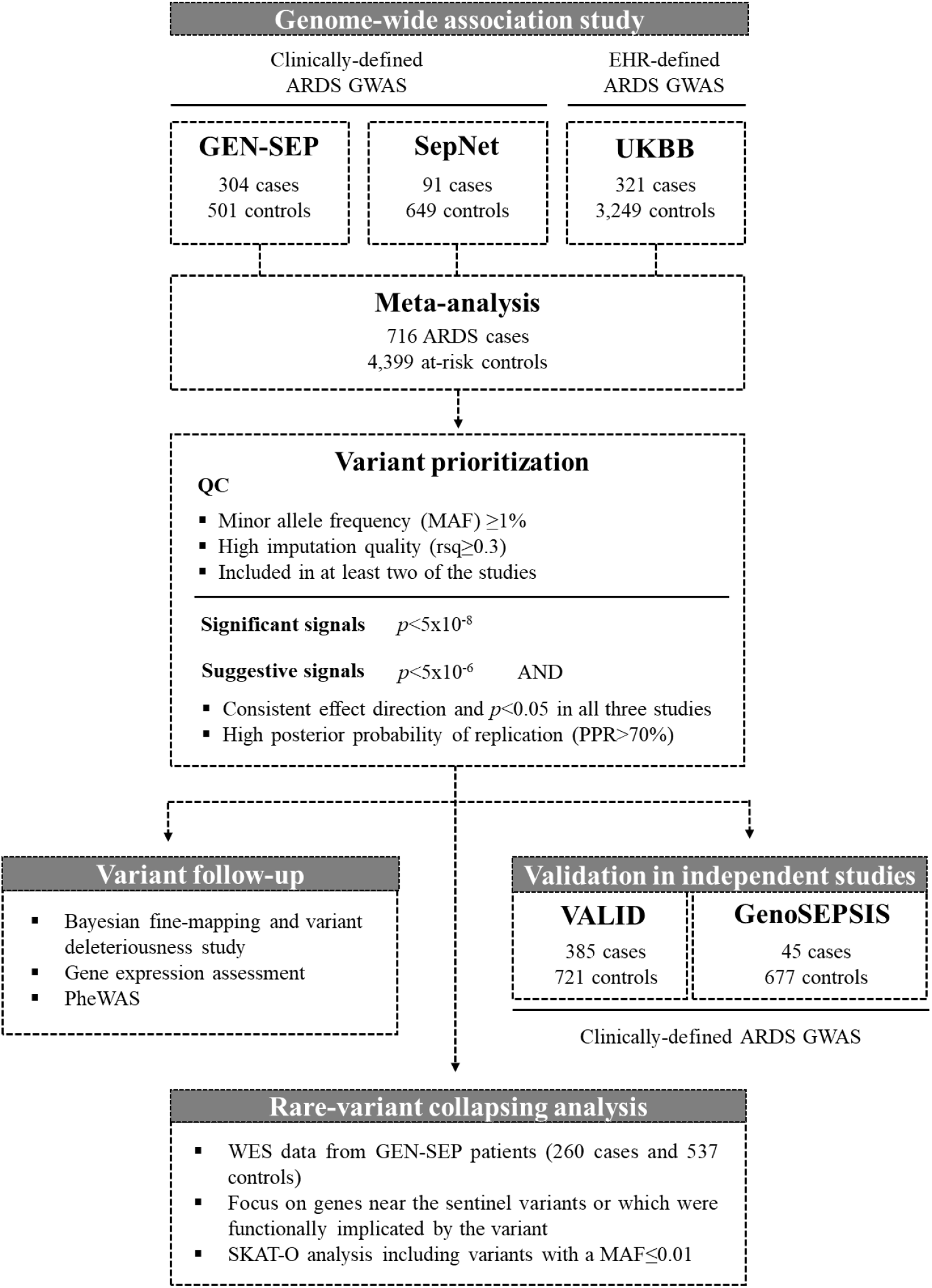
Flowchart of the study design. EHR: Electronic Health Records; PheWAS: Phenome-Wide Association Study; QC: Quality Controls; UKBB: UK Biobank; WES: Whole Exome Sequencing.

The GEN-SEP study was approved by the Ethics Committee from the Hospital Universitario de Canarias (CHUNSC_2018-16). Genetic analyses of SepNet (with separate patient consent for genetic analyses) were integrated in the randomized clinical trials VISEP (recruitment in 2003-2005, registration number: NCT00135473) and MAXSEP (recruitment in 2007-2010, registration number: NCT00534287). All these participating studies were performed according to The Code of Ethics of the World Medical Association (Declaration of Helsinki), and informed consent was obtained from all subjects or their representatives. UK Biobank has approval from the North West Multi-centre Research Ethics Committee (MREC).

### GWAS meta-analysis

A GWAS of ARDS was performed independently in each study. Genotyping, quality control and association testing procedures are described in the appendix. The results of the three GWAS were meta-analysed across the GEN-SEP, SepNet, and UKBB studies using a fixed-effect inverse-variance weighted meta-analysis in METAL (2011 version) (Willer et al., 2010) for all autosomal variants present in at least two studies. The assessment of chromosome X was limited to the GEN-SEP and UKBB studies. To identify independent associated variants, we performed conditional regression analyses using the GCTA COnditional and JOint association analysis using GWAS summary statistics tool (GCTA-COJO) (Zhu et al., 2018) around 1 Mb of the sentinel variant in the prioritised loci, considering the underlying linkage disequilibrium (LD) structure in the study sample. Variant prioritisation was performed based on the replicability of the association signals using the Meta-Analysis Model-Based Assessment of Replicability (MAMBA) (McGuire et al., 2021). MAMBA calculates the posterior probability of replicability (PPR) that a given variant has a non-zero replicable effect across the three studies, indicating the likelihood of a specific genetic variant to replicate.

Genome-wide significant associations were declared for variants satisfying a *p*<5×10^-8^ in the first-stage meta-analysis. We also declared suggestive associations for variants satisfying all the following criteria: *p*<5×10^-6^, consistent effect direction and *p*<0.05 in all studies with available data, and a MAMBA PPR≥70% in the meta-analysis (considered consistent and likely to replicate) (**Figure 1**). Both significant and suggestive loci were prioritised for subsequent analyses. In addition, we also examined our previously reported ARDS protective variant at *FLT1* (rs9508032, chr13:28995940) revealed in the GEN-SEP and SepNet studies (Guillen-Guio et al., 2020) and a variant at *BORCS5* (rs7967111, chr12:12601953) reported in a multi-ancestry GWAS (Du et al., 2021).

### Bayesian fine-mapping and *in-silico* functional assessment

We performed fine-mapping on the association results around the sentinel variants on each prioritised associated locus to identify the credible set of variants that most likely harbours the causal one with 95% confidence. Posterior probabilities (PP) were calculated from approximate Bayes factors for all variants within a region of 1 Mb and a r^2^>0.1 with the sentinel variant (Wakefield, 2009). Variants were considered part of the credible set until their sum of probabilities was >0.95 (see the appendix for details).

To assess the biological consequences of the variants included in the credible sets, we functionally annotated the variants using Ensembl Variant Effect Predictor (VEP) v.105 to obtain the scaled Combined Annotation Dependent Depletion (CADD) score v.1.6 of each variant. Variants with the highest phenotypic impact, based on their CADD score, within each identified credible set were further investigated through additional *in-silico* analyses. We used The Genotype-Tissue Expression (GTEx) Release v.8 data (GTEx Consortium, 2013) to evaluate the existence of expression quantitative trait loci (eQTL) in tissues of interest for the disease (i.e. lung, cultured fibroblasts, whole blood, thyroid, tibial artery, and oesophagus mucosa). We performed colocalisation analysis using the R package *coloc* under a single causal variant assumption (Giambartolomei et al., 2014) to study whether the same causal variant was driving both genetic association with ARDS and gene expression. We also conducted a phenome-wide association study (PheWAS) using publicly available data from Open Targets (Ghoussaini et al., 2021) to identify whether the variants had been previously associated with other phenotypes (*p*<0.005).

### Rare-variant collapsing analysis on prioritised loci

We accessed whole-exome sequencing data from 260 patients with sepsis-associated ARDS and 537 at-risk controls with sepsis from the GEN-SEP cohort (see appendix for more details). Analyses were focused on significant or suggestively associated loci, prioritising genes near the sentinel variants or functionally implicated by the SNP with the highest phenotypic impact. To assess the association of gene-based rare exonic variation, we used the optimal sequence kernel association test (SKAT-O) implemented in EPACTS v.3.6.2 (Lee et al., 2012; EPACTS website). Testing was performed separately for two categories of variants: (i) all variants with MAF≤0.01, and (ii) all MAF≤0.01 variants classified as likely having high phenotypic impact (see appendix). Analyses were controlled for sex, age, and APACHE II, and significance was established at *p*<1.79×10^-3^ considering tests for two categories of variants in 14 genes.

### Validation in independent studies

Prioritised variant associations were assessed in two additional independent studies of patients with sepsis (**Figure 1**). The first study included 1,106 patients of European ancestry (385 sepsis-associated ARDS cases and 721 sepsis controls without ARDS) from the prospective study Validating Acute Lung Injury biomarkers for Diagnosis (VALID) (Siew et al., 2009). Cases and controls were required to have sepsis and organ dysfunction. Diagnosis of ARDS in VALID was done by two-physician review using the Berlin definition. The second study consisted of 45 ARDS cases (Berlin definition) and 677 at-risk controls (primarily with septic shock) from the Genomic Study of Sepsis (GenoSEPSIS) (Martin-Fernandez et al., 2022). Further details of the two studies, including genotyping, imputation and association testing, are described in the appendix.

Both studies were approved by the corresponding Research Ethics Committees (Vanderbilt IRB #051065 for VALID and PI 20-2070 for GenoSEPSIS). Results from the two studies were meta-analysed with METAL following the same procedure as that of the GWAS.

## Results

A total of 716 ARDS cases and 4,399 at-risk controls, and 8,469,305 SNPs with a MAF>0.01, high imputation quality and present in at least two of the three studies were included in the first-stage meta-analysis. There was no evidence of inflation (λ=0.99) (**Figure S1**). A novel variant at chromosome 5q13.3 intergenic to *ANKRD31* and *HMGCR* was genome-wide significantly associated with sepsis-associated ARDS with a high probability of replication (rs116066418, 5q13.3, MAF=0.023, odds ratio (OR)=2.54, 95% confidence interval (CI) =1.83-3.54, *p*=3.43×10^-8^, PPR=0.99) (**Table 1, Figure 2, Figure 3, Figure 4, Figure S2**). Results were robust after excluding non-European individuals (OR(95%CI)=2.57(1.84-3.58), *p*=2.50×10^-8^, PPR=0.99). A further eight loci showed suggestive significance (*p*<5×10^-6^) and high probability of replication (MAMBA PPR≥70%), as well as significance at the nominal level (*p*<0.05) and consistent effect direction in all three studies contributing to the meta-analysis (**Figure S2, Figure S3**). These nine (one genome-wide significant and eight suggestively significant) independent loci were prioritised for further analyses. The association of the variant at *FLT1* identified in our previous GWAS of sepsis-associated ARDS (rs9508032, chr13:28995940), which used data from GEN-SEP and SepNet studies (Guillen-Guio et al., 2020), did not reach significance in the UKBB study (*p*=0.809) (**Figure S4**). The intronic variant at *BORCS5* (rs7967111, chr12:12601953) revealed in the ARDS GWAS performed by Du and colleagues (Du et al., 2021) did not reach significance in our study either (*p*=0.622) (**Figure S4**).

**Table 1.** Association analysis results for the nine signals prioritised after meta-analysis of the three sepsis-associated ARDS studies.

| ID | CHR.<br>band | POS (b37) | Nearest<br>gene/s | NEA/EA | EA_freq | OR (95%CI) | p-value | PPR | Direction* |
| --- | --- | --- | --- | --- | --- | --- | --- | --- | --- |
| rs749263563 | 3p14.2 | 3:58794870 | *FAM3D* | TAGAC/T | 0.023 | 2.66 (1.77, 4.00) | 2.34x10<sup>-6</sup> | 0.96 | ?++ |
| rs183861060 | 4q21.21 | 4:80403211 | *GK2* | A/G | 0.014 | 2.95 (1.93, 4.52) | 6.39x10<sup>-7</sup> | 0.90 | +?+ |
| rs116066418 | 5q13.3 | 5:74588898 | *ANKRD31*<br>, *HMGCR* | T/A | 0.023 | 2.54 (1.83, 3.54) | 3.43x10<sup>-8</sup> | 0.99 | +++ |
| rs59685452 | 6q25.3 | 6:159277046 | *EZR* | G/C | 0.030 | 2.12 (1.58, 2.86) | 6.10x10<sup>-7</sup> | 0.98 | +++ |
| rs12684841 | 9q22.32 | 9:97408971 | *FBP1*,<br>*AOPEP* | A/G | 0.079 | 1.63 (1.34, 1.99) | 1.51x10<sup>-6</sup> | 0.90 | +++ |
| rs11111647 | 12q23.3 | 12:103945224 | *STAB2* | G/A | 0.212 | 1.40 (1.22, 1.62) | 2.97x10<sup>-6</sup> | 0.75 | +++ |
| rs118026254 | 12q24.22 | 12:117415150 | *FBXW8* | G/C | 0.033 | 2.18 (1.60, 2.96) | 8.38x10<sup>-7</sup> | 0.82 | +?+ |
| rs8022645 | 14q23.3 | 14:66327149 | *FUT8*,<br>*CCDC196* | C/T | 0.234 | 1.40 (1.22, 1.61) | 1.28x10<sup>-6</sup> | 0.76 | +++ |
| rs4989808 | 15q26.1 | 15:92466080 | *SLCO3A1* | A/G | 0.020 | 2.66 (1.82, 3.87) | 3.77x10<sup>-7</sup> | 0.94 | +?+ |

CHR: chromosome, CI: confidence interval, EA: effect allele, EA\_freq: frequency of the effect allele, NEA: non-effect allele, OR: Odds ratio, POS: position, PPR: posterior probability of replication (obtained with MAMBA, Meta-Analysis Model-Based Assessment of Replicability).
\*Order for effect Direction is: GEN-SEP – SepNet – UKBB (+ means beta >0 and ? means missing data).

**Figure 2.**
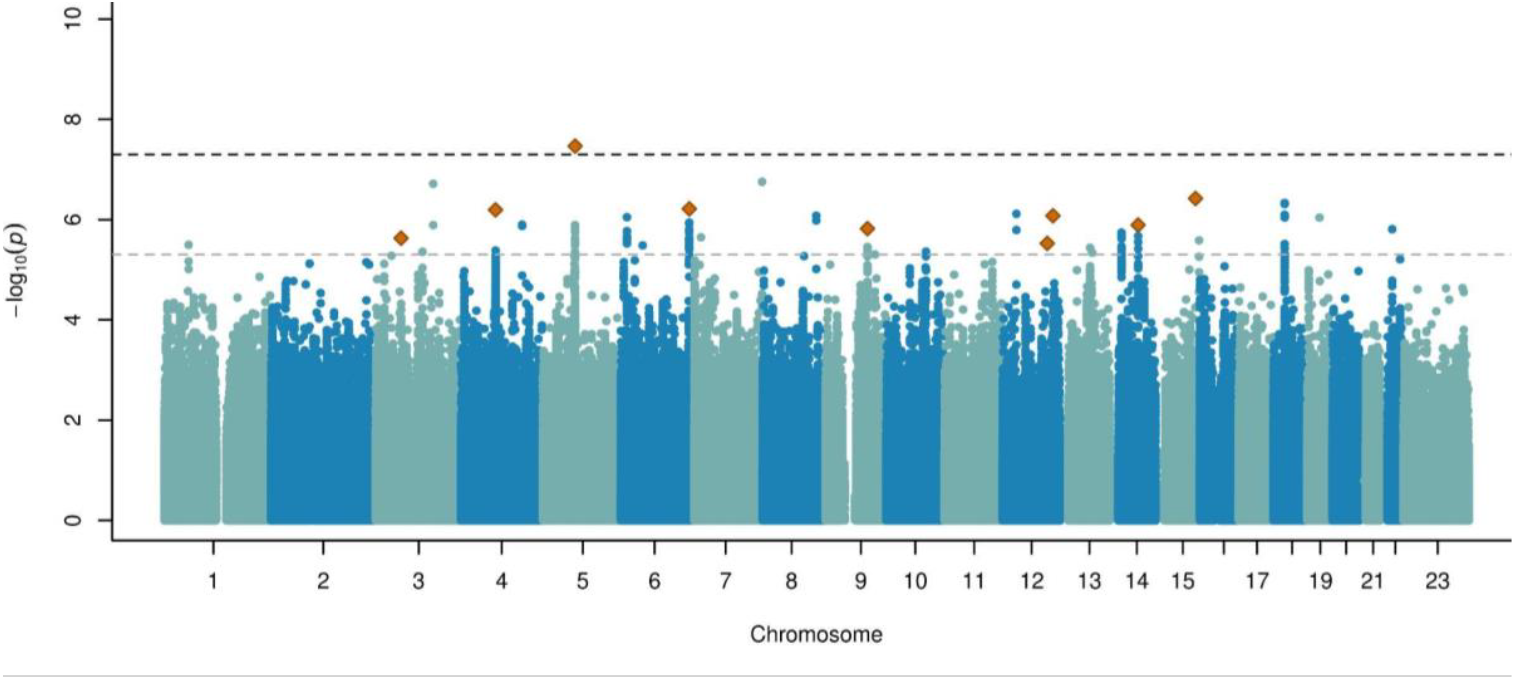
Manhattan plot of the GWAS meta-analysis results. The y-axis shows the transformed *p*-values (-log10 *p*-value), and the x-axis represents the chromosome positions (GRCh37/hg19). Genome-wide significance (*p*-value=5.0×10^-8^) and suggested significance (*p*-value=5.0×10^-6^) thresholds are indicated by the upper and lower horizontal dashed lines, respectively. The X chromosome is represented as 23 and contains the meta-analysis results of GEN-SEP and UKBB. The orange diamond represents the prioritised variants (inflation λ= 0.99).

**Figure 3.**
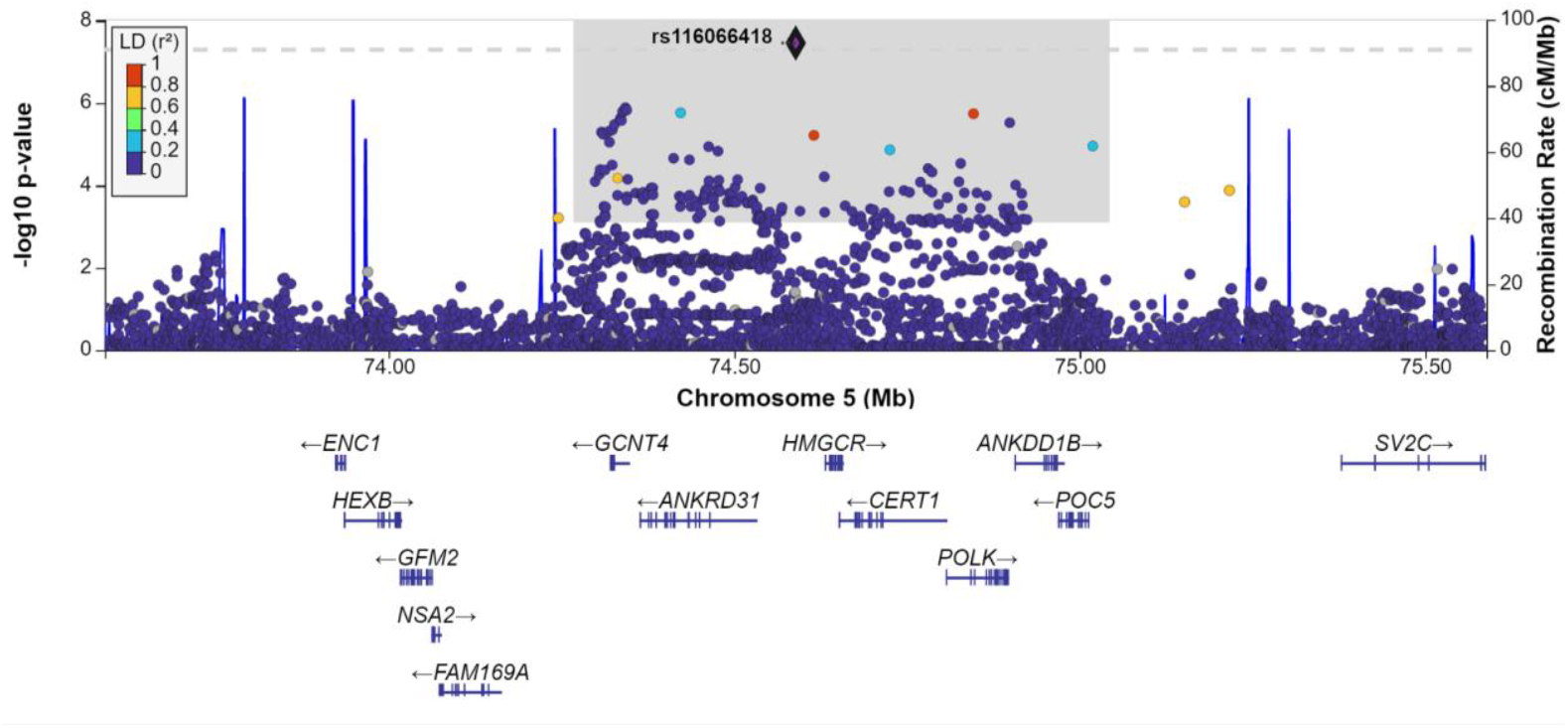
Regional plot of the association results for the genome-wide significant variant at 5q13.3. The y-axis shows the transformed *p*-values (-log10[*p*-value]), and the x-axis represents the chromosome positions (GRCh37/hg19). Genome-wide significance threshold (*p*-value=5.0×10^-8^) is indicated by the horizontal dashed line. Linkage disequilibrium (LD) values (r^2^) are based on the European population data from The 1000 Genomes Project and are represented according to the LD colour scheme of the top left legend. The shaded grey area denotes the variants contained within the credible set. The plot was generated with LocusZoom (http://locuszoom.org/).

**Figure 4.**
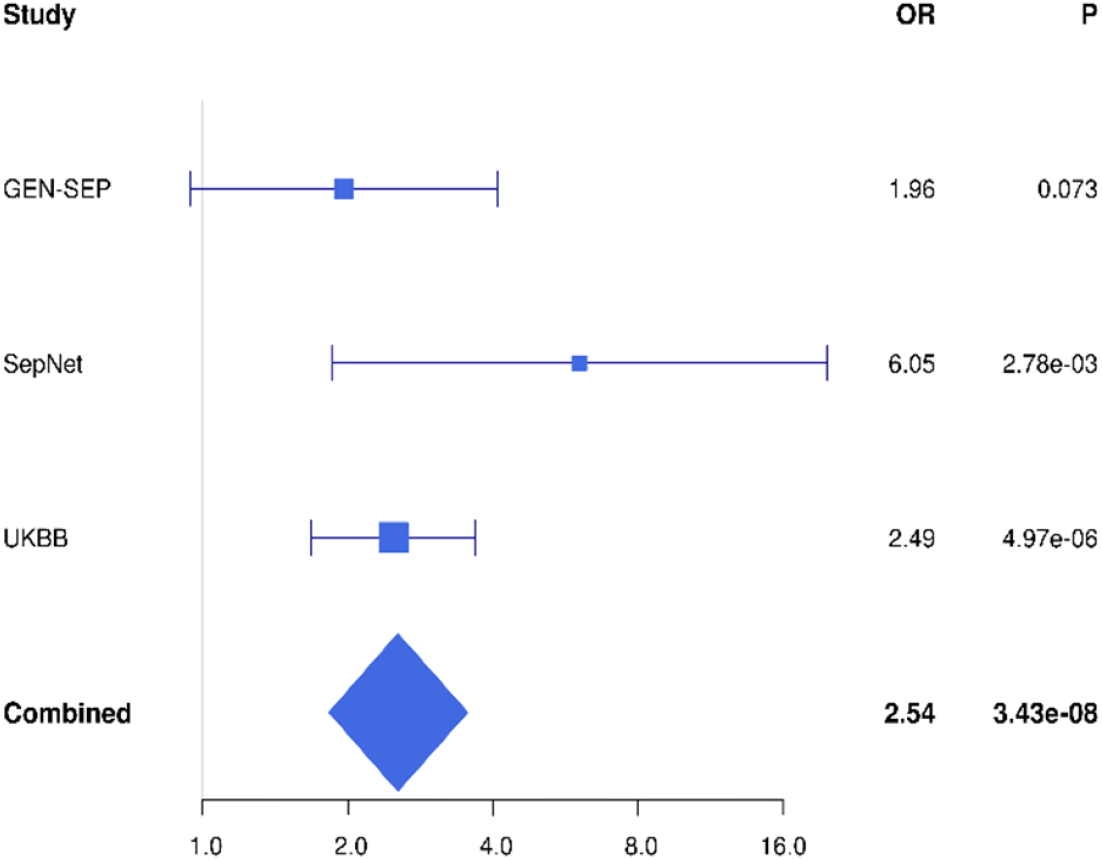
Forest plot of the association results for the genome-wide significant variant at 5q13.3.

Bayesian genetic fine-mapping around each of the nine loci was performed to identify the most likely causal variants driving the association. The 95% credible sets were provided for all the loci except the one at chromosome 15q26.1, due to the absence of LD with the sentinel variant in this region (**Table S3, Table S4, Figure S5**). For each locus, we selected the variant with the highest CADD score, revealing three variants with a high phenotypic impact prediction (CADD>15.00) within the credible sets of the loci at chromosomes 3p14.2 (rs80308704, CADD=17.80), 5q13.3 (rs6893216, CADD=16.66), and 9q22.32 (rs73523269, CADD=15.78) (**Table S3**).

*In-silico* functional evaluation revealed that the risk allele of the 5q13.3 missense variant rs6893216 was associated with increased *POC5* expression in cultured fibroblasts, thyroid and Epstein-Barr Virus-transformed lymphocytes, and with increased *ANKDD1B* expression in artery (**Table S4**). The ARDS GWAS signal colocalised with *ANKDD1B* eQTL signals in tibial artery (colocalisation probability [coloc] >70%). PheWAS results revealed that the risk allele of this variant has also been associated with increased LDL and total cholesterol levels (*p*=7.8×10^-115^ and *p*=4.7×10^-67^, respectively), increased statin medication (*p*=1.8×10^-14^), and higher platelet count (*p*=1.2×10^-8^), amongst others (**Table S4**). The risk allele of the variant at 9q22.32 was associated with reduced expression of *FBP1* in oesophagus, where the causal variant driving ARDS risk and *FBP1* expression was the same (coloc=88.7%). The variant at 3p14.2 did not emerge as a significant eQTL for any genes, and has not been previously associated with other traits at genome-wide level (**Table S4**).

The gene-based rare exonic variant analysis revealed a nominal association (*p*<0.05) between *HMGCR* and ARDS when all variants with MAF<0.01 were considered in the analysis (*p*=0.038), and between *POC5* and sepsis-associated ARDS when all variants with MAF<0.01 and a likely high phenotypic impact were included (*p*=0.011) (**Table 2**). These two genes are near or functionally implicated by the genome wide significant variant at 5q13.3.

**Table 2.** Results of the rare exonic gene-level associations at prioritised loci.

| Locus | Gene* | Chr | Start-End | Test | Variants# | p-value |
| --- | --- | --- | --- | --- | --- | --- |
| 3p14.2 | *FAM3D* | 3 | 58,619,673-58,652,575 | MAF<0.01 | 7 | 0.165 |
|  |  |  |  | MAF<0.01 \| high-impact | 5 | 0.068 |
| 4q21.21 | *GK2* | 4 | 80,327,508-80,329,372 | MAF<0.01 | 11 | 0.105 |
|  |  |  |  | MAF<0.01 \| high-impact | 9 | 0.128 |
| 5q13.3 | *ANKRD31* | 5 | 74,364,100-74,532,703 | MAF<0.01 | 27 | 1.000 |
|  |  |  |  | MAF<0.01 \| high-impact | 15 | 1.000 |
|  | *HMGCR* | 5 | 74,632,154-74,657,929 | MAF<0.01 | 9 | **0.038** |
|  |  |  |  | MAF<0.01 \| high-impact | 7 | 0.142 |
|  | *ANKDD1B* | 5 | 74,907,284-74,967,671 | MAF<0.01 | 10 | 0.125 |
|  |  |  |  | MAF<0.01 \| high-impact | 2 | 0.145 |
|  | *POC5* | 5 | 74,969,949-75,013,313 | MAF<0.01 | 9 | 0.163 |
|  |  |  |  | MAF<0.01 \| high-impact | 8 | **0.011** |
| 6q25.3 | *EZR* | 6 | 159,186,773-159,240,444 | MAF<0.01 | 7 | 0.714 |
|  |  |  |  | MAF<0.01 \| high-impact | 6 | 0.329 |
| 9q22.32 | *MFSD14B* | 9 | 97,136,833-97,223,324 | MAF<0.01 | 8 | 0.688 |
|  |  |  |  | MAF<0.01 \| high-impact | 6 | 0.579 |
|  | *FBP1* | 9 | 97,365,415-97,402,531 | MAF<0.01 | 3 | 0.298 |
|  |  |  |  | MAF<0.01 \| high-impact | 3 | 0.289 |
|  | *AOPEP* | 9 | 97,488,983-97,849,441 | MAF<0.01 | 17 | 0.306 |
|  |  |  |  | MAF<0.01 \| high-impact | 13 | 0.109 |
| 12q23.3 | *STAB2* | 12 | 103,981,051-104,160,505 | MAF<0.01 | 59 | 0.879 |
|  |  |  |  | MAF<0.01 \| high-impact | 52 | 0.915 |
| 12q24.22 | *FBXW8* | 12 | 117,348,761-117,468,953 | MAF<0.01 | 17 | 0.778 |
|  |  |  |  | MAF<0.01 \| high-impact | 14 | 0.727 |
| 14q23.3 | *FUT8* | 14 | 65,877,310-66,210,839 | MAF<0.01 | 4 | 0.383 |
|  |  |  |  | MAF<0.01 \| high-impact | 4 | 0.384 |
| 15q26.1 | *SLCO3A1* | 15 | 92,396,925-92,715,665 | MAF<0.01 | 6 | 0.071 |
|  |  |  |  | MAF<0.01 \| high-impact | 5 | 0.127 |

Chr: Chromosome; MAF: Minor Allele Frequency. Gene positions based on GRCh37/hg19, Ensembl. \*Genes near the sentinel variants or which were functionally implicated by the highest phenotypic impact SNP in each locus. #Number of variants tested. Results were not available for *CCDC196*.

Finally, the leading independent variants from the nine prioritised loci were tested for association with sepsis-associated ARDS in VALID and GenoSEPSIS cohorts. While none of the variants reached nominal significance (*p*<0.05) in either independent replication cohorts, rs11111647 (12q23.3) became slightly more significant after the second-stage meta-analysis, and three variants had consistent direction of effects across the five studies, including the genome-wide significant variant at 5q13.3 and variants located at the 6q25.3 and 12q23.3 loci (**Table 3**).

**Table 3.** Association results in the validation cohorts.

|  |  | First-stage Meta-analysis<br>(716 cases/4,399 controls) |  | VALID<br>(385 cases/721 controls) |  | GenoSEPSIS (45<br>cases/677 controls) |  | Second-stage meta-analysis<br>(1,146 cases/5,797 controls) |  |
| --- | --- | --- | --- | --- | --- | --- | --- | --- | --- |
| ID<br>(POS b37) | NEA/E<br>A | OR<br>(95%CI) | *p*-value | OR<br>(95%CI) | *p*-value | OR<br>(95%CI) | *p*-value | OR<br>(95%CI) | *p*-value |
| **rs749263563**<br>(3:58794870,<br>3p14.2) | TAGA<br>C/T | 2.66<br>(1.77, 4.00) | 2.34x10<sup>-6</sup> | 0.85<br>(0.48, 1.53) | 0.593 | NA | NA | 1.54<br>(0.50, 4.69) | 4.12x10<sup>-5</sup> |
| **rs183861060**<br>(4:80403211,<br>4q21.21) | A/G | 2.95<br>(1.93, 4.52) | 6.39x10<sup>-7</sup> | 1.08<br>(0.69, 1.69) | 0.749 | 0.78<br>(0.18, 3.45) | 0.740 | 1.52<br>(0.65, 3.58) | 2.02x10<sup>-5</sup> |
| **rs116066418**<br>(5:74588898,<br>5q13.3) | T/A | 2.54<br>(1.83, 3.54) | 3.43x10<sup>-8</sup> | 1.28<br>(0.73, 2.25) | 0.391 | 1.22<br>(0.26, 5.65) | 0.802 | 2.09<br>(1.58, 2.77) | 2.48x10<sup>-7</sup> |
| **rs59685452**<br>(6:159277046<br>, 6q25.3) | G/C | 2.12<br>(1.58, 2.86) | 6.10x10<sup>-7</sup> | 1.01<br>(0.55, 1.85) | 0.972 | 1.78<br>(0.66, 4.80) | 0.255 | 1.84<br>(1.42, 2.38) | 3.38x10<sup>-6</sup> |
| **rs12684841**<br>(9:97408971,<br>9q22.32) | A/G | 1.63<br>(1.34, 1.99) | 1.51x10<sup>-6</sup> | 0.97<br>(0.70, 1.34) | 0.846 | 1.46<br>(0.71, 3.00) | 0.300 | 1.32<br>(0.89, 1.95) | 1.76x10<sup>-5</sup> |
| **rs11111647**<br>(12:10394522<br>4, 12q23.3) | G/A | 1.40<br>(1.22, 1.62) | 2.97x10<sup>-6</sup> | 1.10<br>(0.89, 1.36) | 0.378 | 1.62<br>(1.00, 2.65) | 0.051 | 1.32<br>(1.17, 1.48) | 2.91x10<sup>-6</sup> |
| **rs118026254**<br>(12:11741515<br>0, 12q24.22) | G/C | 2.18<br>(1.60, 2.96) | 8.38x10<sup>-7</sup> | 1.14<br>(0.71, 1.83) | 0.596 | 0.41<br>(0.09, 1.80) | 0.239 | 1.30<br>(0.65, 2.60) | 1.80x10<sup>-5</sup> |
| **rs8022645**<br>(14:66327149<br>, 14q23.3) | C/T | 1.40<br>(1.22, 1.61) | 1.28x10<sup>-6</sup> | 0.97<br>(0.79, 1.20) | 0.792 | 1.05<br>(0.63, 1.76) | 0.844 | 1.16<br>(0.86, 1.55) | 2.77x10<sup>-5</sup> |
| **rs4989808**<br>(15:92466080<br>, 15q26.1) | A/G | 2.66<br>(1.82, 3.87) | 3.77x10<sup>-7</sup> | 1.01<br>(0.59, 1.73) | 0.967 | 0.66<br>(0.08, 5.28) | 0.698 | 1.49<br>(0.63, 3.50) | 8.98x10<sup>-6</sup> |

CI: confidence interval, EA: effect allele, NEA: non-effect allele, OR: Odds ratio, POS: chromosome position.

## Discussion

To our knowledge, we have performed the largest GWAS of sepsis-associated ARDS to date. Our findings have revealed a novel genome-wide common genetic variant associated with sepsis-associated ARDS risk, as well as eight additional loci with internal evidence of replicability. Although none of the signals were validated in two additional studies, all nine signals satisfied the stringent internal replication criteria.

The 5q13.3 locus reaching genome-wide significance has been previously associated with increased cholesterol levels [Open targets, (Klimentidis et al., 2020)]. The most significant variant was located in the intergenic region of *ANKRD31* and *HMGCR*. The risk allele of the highest phenotypic impact variant at this locus colocalised with increased expression of *ANKRD31* in tibial artery, while our gene-based rare exonic variant analysis further supported the role of *HMGCR* in sepsis-associated ARDS, highlighting the potential relevance of these genes in ARDS. *ANKRD31* encodes the Ankyrin Repeat Domain-Containing Protein 31, involved in protein-protein interactions, while *HMGCR* encodes the 3-Hydroxy-3-Methylglutaryl-CoA (HMG-CoA) Reductase, the rate-limiting enzyme for cholesterol synthesis. HMG-CoA reductase inhibitors (statins) have been proposed as treatment for ARDS (McAuley et al., 2014). This is supported by several studies in animal and cellular models that reported simvastatin as a protective treatment from lung injury by maintaining the alveolar-capillary barrier integrity and reduce inflammation (Terblanche et al., 2006; Singla and Jacobson, 2012). However, a number of clinical trials have shown no improvement in sepsis or ARDS outcomes by simvastatin (Adhikari et al., 2004; McAuley et al., 2018; Hofmaenner et al., 2022). Recent studies support that ARDS patients with a hyperinflammatory phenotype could have a survival benefit from simvastatin treatment (Calfee et al., 2018). Additionally, a recent multicentre trial study suggested that low cholesterol patients had lower mortality when simvastatin was used, encouraging future studies evaluating this drug in this group of patients (Pienkos et al., 2023). A computational analysis of omics data in ARDS also supported the cholesterol metabolism dysregulation as one of the main landmarks underlying ARDS pathobiology (Millar et al., 2024). All of this would suggest that the use of statins may be beneficial when used for particular ARDS endotypes.

The 9q22.32 variant associated with ARDS risk, with a high predicted phenotypic impact, was implicated in decreased *FBP1* expression in oesophagus mucosa and pituitary. *FBP1* encodes the fructose-bisphosphatase 1, which is a regulator of appetite and adiposity and whose deficiency is associated with hypoglycaemia (Bijarnia-Mahay et al., 1993; Liang et al., 2023). *FBP1* has been found to be dysregulated under some viral infections, including SARS-CoV-2 infections (Zhang et al., 2021; Sengupta et al., 2022). Moreover, Bhargava and colleagues reported that *FBP1* was differentially expressed in bronchoalveolar lavage fluid from ARDS survivors when compared with non-survivors (Bhargava et al., 2017).

We acknowledge some strengths and limitations of this study. Among the strengths of this study, we aimed to address the heterogeneity of the syndrome by comparing ARDS cases with at-risk controls (mainly with sepsis) who did not develop ARDS. This allowed us to identify robust and concordant associations across studies from three different countries, involving different health care systems. Furthermore, we provided further analyses, including gene expression assessment and a rare variant collapsing approach, to support our meta-GWAS findings. A main limitation is the reduced number of patients of non-European genetic ancestry, which limits the ability to evaluate the generalisability of findings to other populations. Additionally, while we used MAMBA to prioritise loci for follow-up analyses, this method is designed for larges datasets and may not perform optimally with our limited dataset. In fact, the follow-up of the six prioritised variants in two additional studies did not evidence significant findings, although three of the prioritised variants (including the genome-wide significant locus) had the same direction of effect in all the five studies and one became slightly more significant when the results were meta-analysed. The lack of validation in the GenoSEPSIS study could be due to the limited sample size and subsequent reduced statistical power. The UKBB study’s reliance on electronic health records for case and control selection may result in less thoroughly characterised phenotypes compared to other cohorts, including the absence of APACHE scores for model adjustment, potentially introducing misclassification bias into the results. This may provide an explanation for the non-replication of the variant association at *FLT1* reported in a previous sepsis-associated ARDS GWAS (Guillen-Guio et al., 2020) within the UKBB study.

In summary, we describe the results of the largest and more extensive GWAS of sepsis-associated ARDS, reporting a novel common variant association with ARDS susceptibility previously linked to cholesterol levels. The reported locus and subsequent rare-variant analysis suggest that *HMGCR* could be important in the pathophysiology of the syndrome. These findings, coupled with the accumulated evidence, underscore the potential of the lipid metabolism pathway as a target for ARDS treatment. Additional independent replication of these signals and further functional evidence is needed to validate our results.

## Statements

### Funding

BGG is supported by Wellcome Trust (grant 221680/Z/20/Z). For the purpose of open access, the author has applied a CC BY public copyright license to any Author Accepted Manuscript version arising from this submission. ESP and ETH are funded by Gobierno de Canarias and Social European Fund “Canarias Avanza con Europa” (grants TESIS2022010042 and TESIS2021010046 respectively) and Social European Fund Plus (EST2023010007). VEK is funded by National Heart, Lung, and Blood Institute (grant K01HL157755) and ATS Research Program. AS was supported by the German Ministry of Education and Research (01 ZZ 1803C) within the Smart Medical Information Technology for Healthcare consortium. LVW holds a GlaxoSmithKline Asthma + Lung UK Chair in Respiratory Research (C17-1). JV was funded by Instituto de Salud Carlos III, Madrid, Spain (PI19/00141, AC21_2/00039), ERAPerMed (JTC_2021), the European Regional Development Funds, Fundación Canaria Instituto de Investigación Sanitaria de Canarias. CF is funded by Instituto de Salud Carlos III (PI1400844, PI17/00610, PI20/00876 and PI23/00980); European Union (ERDF) ‘A way of making Europe’, Agencia Estatal de Investigación (RTC-2017-6471-1), ITER agreements (OA17/008 and OA23/043), and Cabildo Insular de Tenerife (CGIEU0000219140 and ‘Apuestas científicas del ITER para colaborar en la lucha contra la COVID-19’). The SepNet GWAS project was supported by the Paul-Martini-Sepsis Research Group (funded by the Thuringian Ministry of Education, Science, and Culture [ProExcellence; grant PE 108–2]), the Thuringian Foundation for Technology, Innovation, and Research, the German Sepsis Society, and the Jena Center of Sepsis Control and Care (funded by the German Ministry of Education and Research [01 EO 1002 and 01 EO 1502]). The VISEP and MAXSEP trials from the SepNet Study Group were supported by a German Ministry of Education and Research grant (01 KI 0106), and by unrestricted grants from B Braun, HemoCue, Novo Nordisk, AstraZeneca (Wedel, Germany), and Bayer HealthCare (Leverkusen, Germany). This research was partially supported by the National Institute for Health Research (NIHR) Leicester Biomedical Research Centre. This research includes use of the UK Biobank through application 648 and used the ALICE High Performance Computing Facility at the University of Leicester.

### Competing interests

DC and BLY are full-time employees of Genentech. BLY holds stock options in Roche. MS receives funding from Pfizer for a project not related to this research. LVW reports research funding from GlaxoSmithKline, Genentech and Orion Pharma, and consultancy for Galapagos and GlaxoSmithKline, outside of the submitted work. The other authors declare no competing interests.

### Authors contribution

BG-G, ES-P, ET-H, TH-B, JML-S, DC, RG-M, LAR-R, OCL, RJA, MS, and AS performed the analyses. AC, RC, MB-C, AC, ET, VEK, LBW, BLY, MS, AS, JV, LVW, and CF participated in data collection. LVW and CF supervised the study. BG-G, LVW, and CF obtained funding. BG-G, ES-P, and CF wrote the first draft of the manuscript. All authors revised and approved the final version of the manuscript.

### Data availability

All data produced in the present study are available upon reasonable request to the authors

