## appendix for "Genome-wide association study of susceptibility to acute respiratory distress syndrome"

*Equal contribution as senior authors

### Supplementary Methods

#### Sample description

We performed a GWAS meta-analysis on 5,115 patients with sepsis from three independent studies (**Figure 1**): the GENetics of SEPsis-induced ARDS Network (GEN-SEP), The Critical Care Trials Group of the German Sepsis Competence Network (SepNet), and UK Biobank (UKBB). Cases were defined as critical patients who developed ARDS, mainly associated with all-cause sepsis, while the majority of controls comprised critically ill patients with sepsis who did not develop ARDS. The study comprised a total of 716 sepsis-associated ARDS cases and 4,399 sepsis controls, including 805 patients from the GEN-SEP cohort (304 cases and 501 at-risk controls), 740 from SepNet (91 cases and 649 at-risk controls), and 3,570 from UKBB (321 cases and 3,249 at-risk controls).

In GEN-SEP and SepNet, patients with a diagnosis of sepsis (Singer et al., 2016) admitted to the Intensive Care Units (ICUs) were followed for ARDS development according to Berlin definition criteria (Force et al., 2012). GEN-SEP is a multicentre, observational study conducted in a Spanish network of ICUs comprising adult patients with all-cause sepsis of European ancestry recruited in two periods (in 2002-2015 and in 2016-2019) (Hernandez-Beeftink et al., 2022). The GEN-SEP study was approved by the Ethics Committee from the Hospital Universitario de Canarias (CHUNSC_2018-16). SepNet is integrated by the VISEP (recruitment in 2003-2005, registration number: NCT00135473) and MAXSEP (recruitment in 2007-2010, registration number: NCT00534287) randomised controlled trials and includes European ancestry patients with all-cause sepsis from German hospitals (Scherag et al., 2016).

For the UKBB cohort, ARDS cases were obtained from UK Biobank based on the International Classification of Diseases 10th Revision (ICD-10) diagnosis code J80 (“Adult respiratory distress syndrome”). Of these, 75.4% ARDS cases were associated with pulmonary or non-pulmonary septicaemia, including pneumonia. The remaining 24.6% ARDS cases were associated with infections due to complications of surgical and medical care (2.18%), pancreatitis (1.56%), major injury (1.56%), inhalation of harmful substances (0.93%), drug poisoning (0.62%), and unspecified “other causes” (17.76%). The inclusion of non-septicaemia patients aims to enhance the statistical power of the study. The list of ICD-10 codes used to determine the ARDS development causes is provided in **Table S1**. More than 92% of individuals were of European ancestry (**Table S2**). Controls in the UKBB study comprised critically ill patients matched for ARDS development cause (i.e., 75.4% pneumonia and/or septicaemia, 24.6% other causes), sex, age, and ancestry. UK Biobank has approval from the North West Multi-centre Research Ethics Committee (MREC).

#### Genotyping, imputation and association studies

In GEN-SEP, single nucleotide polymorphism (SNP) genotyping was performed using the Axiom Genome-Wide Human CEU 1 array (Thermo Fisher Scientific) in the National Genotyping Center (CeGen), Universidad de Santiago de Compostela Node, Spain. Variant calling was done with AffyPipe v.2.10.0, following the manufacturer recommendations. Genotyping quality control was conducted using PLINK v.1.9 and R v.3.6.0 in order to exclude patients with missing clinical information, call rates (CR) <95%, sex discordance, high kinship degree (PIHAT>0.2), or heterozygosity outliers. Those variants with CR<95%, minor allele frequency (MAF) <1%, or that strongly deviated from the Hardy Weinberg equilibrium (HWE) expectations ($p<1.0\text{x}10^{-6}$) were excluded from the analyses. We performed a principal component (PC) analysis (PCA) based on >114,000 independent SNPs to identify and exclude ancestry outliers. Variant imputation was performed with the Michigan Imputation Server using the Haplotype Reference Consortium (HRC) panel v.1.1 (Loh et al., 2016). For variants with MAF>1% and high imputation quality (Rsq>0.3), we performed genetic association testing with

EPACTS v.3.2.6 (EPACTS website) using a logistic regression model. We adjusted for sex, age, and the Acute Physiology and Chronic Health Evaluation II (APACHE II) score as described in our prior study (Guillen-Guio et al., 2020). No PCs were included in the model due to the absence of substructure in this cohort. For the X chromosome, we followed the quality controls described above, separately for males and females. In addition, a SNP filtering was performed based on the HWE deviations calculated from females, and results for males and females were then meta-analysed using METASOFT (Han and Eskin, 2011).

In SepNet, SNP genotyping was obtained with the HumanOmniExpressExome array (Illumina, Inc.). Those patients without discordance between the declared and genetic sex, CR<98%, implausible heterozygosity (<20% and >26%) or that were detected as genetic outliers based on PCA were excluded. Variants with a CR≤95%, MAF<1%, or strongly deviating from HWE ($p$<1.0x10$^{-6}$) were also removed from the analyses. Variant imputation was obtained in prephased data from SHAPEIT v.2.r790 (Delaneau et al., 2012) using IMPUTE2 v.2.3.0 (Howie et al., 2009) and the 1000 Genomes Project data (phase1, v3) as a reference panel (The 1000 Genomes Project Consortium, 2015). Logistic regression was used for genetic association testing with SNPTEST v.2.5 (Marchini and Howie, 2010) for variants with MAF>1%, HWE $p$-value>1.0x10$^{-10}$, and high imputation quality (INFO>0.8). The first three genetic PCs, sex, age, and APACHE II were included in the model as covariates. Only autosomes were evaluated in this study.

In UKBB, imputed genetic data and phenotypes were obtained through UK Biobank Application 648 (Bycroft et al., 2018). Genotyping was done using the UK BiLEVE and Applied Biosystems UK Biobank Axiom Arrays (Thermo Fisher Scientific). Individuals were removed if they had a final individual CR<95%, discordance between recorded and genetically inferred sex, were related (Kinship≥0.15), or if they had an extreme PC-adjusted heterozygosity rate. Ethnic groups were defined through K-means clustering using the first two PCs, as previously described (Shrine et al., 2019). Only variants included on both the UK BiLEVE and UK Biobank Axiom arrays, and satisfying a HWE p>1.0x10$^{-6}$, CR≥90%, and MAF≥ 0.0001 were considered. HRC served as the primary reference panel for variant imputation (Bycroft et al., 2018). Additionally, imputation was conducted with the combined UK10K and 1000 Genomes phase 3 reference panels, integrating the results with those from HRC. Quality controls were consistently applied to chromosome X. HWE deviation analysis was conducted exclusively on females on the sex-specific region of the X chromosome. Logistic regression analyses were done using PLINK v2.00 (Chang et al., 2015) and models were adjusted for the first ten genetic PCs, sex, and age. Only variants with MAF>1% and an imputation quality score ≥0.3 were considered in the analyses.

#### Bayesian fine-mapping

We performed a Bayesian fine mapping on the association results around 1 Mb of each sentinel variant (significantly or suggestively associated), considering all with a r$^2$>0.1 with the sentinel variant. The posterior probabilities (PPs) were calculated from the approximate Bayes factors (ABFs) using the formula proposed by Wakefield (Wakefield, 2009):

$$\boldsymbol{ABF} = \frac{\mathbf{1}}{\sqrt{\mathbf{1} - \frac{\boldsymbol{W}}{\boldsymbol{V} + \boldsymbol{w}}}} \boldsymbol{exp}\left(-\frac{\boldsymbol{Z}^{\mathbf{2}}\,\boldsymbol{W}}{\mathbf{2}\boldsymbol{V} + \boldsymbol{W}}\right)$$

Where, for each variant, V is the variance of the effect size, W is the Wakefield prior of 0.4, equivalent to a 95% confidence, and Z is the Z-score.

The approximate posterior probability for each variant was set as its ABF divided by the sum of the ABFs of all variants in the locus. Variants were sorted into descending order of their computed posterior probability and then cumulatively summed until the credible set threshold was exceeded (>0.95).

#### Whole-exome sequencing, bioinformatics processing, and variant annotation

Whole-exome sequencing was available from 260 patients with sepsis-associated ARDS and 537 at-risk controls with sepsis from the GEN-SEP study.

Libraries were prepared using the DNA Prep with Enrichment kit (Illumina Inc.) or SureSelect XT HS2 DNA (Agilent Technologies Inc.) as detailed elsewhere (Diaz-de Usera et al., 2020). Library sizes and concentration were obtained with a TapeStation 4200 (Agilent Technologies Inc.) and the Qubit dsDNA HS Assay (Thermo Fisher Scientific). Sequencing was conducted on Illumina NextSeq550, HiSeq4000, or NovaSeq6000 using 75 or 100 base paired-end reads (depending on the instrument) to an average of 100X depth and including 1% of PhiX control V3 (Illumina Inc.). Sequencing was done at Instituto Tecnológico y de Energías Renovables (ITER, Santa Cruz de Tenerife, Spain).

Pre-processing of sequencing data was conducted by bcl2fastq v.2.18 for demultiplexing and BWA-MEM v.0.7.15 (Li and Durbin, 2009) to align reads to the GRCh37/hg19 reference. Resulting BAM files were assessed using SAMtools v.1.3 (Danecek et al., 2021) and Picard v.2.10.10 (Picard website). Calling of small germline variants (SNPs and indels of up to 50 bp) was performed using GATK HaplotypeCaller v.3.8 (Poplin et al., 2018) with a padding of 100 bp around the capture targets and following the GATK Best Practices workflow recommendations. Variants were filtered using BCFtools v.1.16 (Danecek et al., 2021) based on "PASS" filter, missingness (FMISS) $\leq 0.05$, genotype quality (GQ) $\geq 20$, and depth of coverage (DP) $\geq 10$. Functional annotation of variants was performed on the resulting filtered callsets located on the GWAS-prioritised genes. Variants were annotated for population allele frequency, variant type, protein function, and pathogenic potential with Ensembl Variant Effect Predictor (VEP) v.105 and ANNOVAR v.07.06.20 (Wang et al., 2010) based on different databases. Analyses were conducted at the Teide-HPC supercomputing facility (http://teidehpc.iter.es/en).

#### Rare exonic variant association testing

To assess the association of gene-based rare exonic variation with SKAT-O, we selected the genes that were closest to the prioritised variant and those with the highest functional relevance in the colocalisation. We performed these tests including only the variants with MAF$\leq$0.01 (i.e., with MAF$\leq$0.01 in The 1000 Genomes Project Phase 3 or in gnomAD v.2.1.1, or that were absent from these sources but present in the GEN-SEP patients) or only the variants that had MAF$\leq$0.01 and considered of likely high biological impact based on scaled CADD score > Mutation Significance Cutoff (MSC) or with CADD >15 in absence of MSC information for the given gene.

#### Validation cohorts

The Validating Acute Lung Injury biomarkers for Diagnosis (VALID) study comprised 1,106 patients with sepsis (Siew et al., 2009). The criterion for classifying a patient as a case (n=385) was that the patient had Berlin definition of ARDS on two consecutive days and was on mechanical ventilation for at least one day. At-risk controls (n=721) were those ICU patients with sepsis participating in the trial without ARDS at enrolment or for the next three days of follow-up. SNP genotyping was done with a custom Global Screening Array (Illumina Inc.) and variants were imputed using a reference panel consisting of Genentech's whole-genome sequencing data plus The 1000 Genomes Project dataset (all at a depth of 30X) (Sudmant et al., 2015). Variant imputation was done using BEAGLE v.5.1 (Browning et al., 2018) and association testing was carried out using logistic regression models in PLINK v.1.9 (Purcell et al., 2007), correcting for genetically inferred sex, age, and first five PCs. All individuals included in the analysis had ≥70% European genetic ancestry according to ADMIXTURE v.1.3 (Alexander et al., 2009).

The Genomic Study of Sepsis (GenoSEPSIS) cohort consisted of 722 Spanish ICU patients recruited in the Hospital Clinico Universitario de Valladolid (Spain) (Martin-Fernandez et al., 2022). This study included 45 ARDS cases according to Berlin definition criteria (Force et al., 2012) and 677 at-risk controls, mostly with septic shock. SNP genotyping was done with the Spain Biobank Array (Thermo Fisher Scientific) in the National Genotyping Center (CeGen), Universidad de Santiago de Compostela Node, Spain. Quality control, filtering, imputation, and association testing were conducted following the same procedures as in the GEN-SEP study and adjusting for the first four PCs and sex.

Both studies were approved by the relevant Research Ethics Committees (Vanderbilt IRB #051065 for VALID and PI 20-2070 for GenoSEPSIS).

Shrine, N., Guyatt, A. L., Erzurumluoglu, A. M., Jackson, V. E., Hobbs, B. D., Melbourne, C. A., Batini, C., Fawcett, K. A., Song, K., Sakornsakolpat, P., Li, X., Boxall, R., Reeve, N. F., Obeidat, M., Zhao, J. H., Wielscher, M., Weiss, S., Kentistou, K. A., Cook, J. P., Sun, B. B., Zhou, J., Hui, J., Karrasch, S., Imboden, M., Harris, S. E., Marten, J., Enroth, S., Kerr, S. M., Surakka, I., Vitart, V., Lehtimaki, T., Allen, R. J., Bakke, P. S., Beaty, T. H., Bleecker, E. R., Bosse, Y., Brandsma, C. A., Chen, Z., Crapo, J. D., Danesh, J., DeMeo, D. L., Dudbridge, F., Ewert, R., Gieger, C., Gulsvik, A., Hansell, A. L., Hao, K., Hoffman, J. D., Hokanson, J. E., Homuth, G., Joshi, P. K., Joubert, P., Langenberg, C., Li, X., Li, L., Lin, K., Lind, L., Locantore, N., Luan, J., Mahajan, A., Maranville, J. C., Murray, A., Nickle, D. C., Packer, R., Parker, M. M., Paynton, M. L., Porteous, D. J., Prokopenko, D., Qiao, D., Rawal, R., Runz, H., Sayers, I., Sin, D. D., Smith, B. H., Soler Artigas, M., Sparrow, D., Tal-Singer, R., Timmers, P., Van den

Berge, M., Whittaker, J. C., Woodruff, P. G., Yerges-Armstrong, L. M., Troyanskaya, O. G., Raitakari, O. T., Kahonen, M., Polasek, O., Gyllensten, U., Rudan, I., Deary, I. J., Probst-Hensch, N. M., Schulz, H., James, A. L., Wilson, J. F., Stubbe, B., Zeggini, E., Jarvelin, M. R., Wareham, N., Silverman, E. K., Hayward, C., Morris, A. P., et al. 2019. New genetic signals for lung function highlight pathways and chronic obstructive pulmonary disease associations across multiple ancestries. *Nat Genet,* 51(3)**,** pp 481-493.

Siew, E. D., Ware, L. B., Gebretsadik, T., Shintani, A., Moons, K. G., Wickersham, N., Bossert, F. & Ikizler, T. A. 2009. Urine neutrophil gelatinase-associated lipocalin moderately predicts acute kidney injury in critically ill adults. *J Am Soc Nephrol,* 20(8)**,** pp 1823-32.

Singer, M., Deutschman, C. S., Seymour, C. W., Shankar-Hari, M., Annane, D., Bauer, M., Bellomo, R., Bernard, G. R., Chiche, J. D., Coopersmith, C. M., Hotchkiss, R. S., Levy, M. M., Marshall, J. C., Martin, G. S., Opal, S. M., Rubenfeld, G. D., van der Poll, T., Vincent, J. L. & Angus, D. C. 2016. The Third International Consensus Definitions for Sepsis and Septic Shock (Sepsis-3). *JAMA,* 315(8)**,** pp 801-10.

Sudmant, P. H., Rausch, T., Gardner, E. J., Handsaker, R. E., Abyzov, A., Huddleston, J., Zhang, Y., Ye, K., Jun, G., Fritz, M. H., Konkel, M. K., Malhotra, A., Stutz, A. M., Shi, X., Casale, F. P., Chen, J., Hormozdiari, F., Dayama, G., Chen, K., Malig, M., Chaisson, M. J. P., Walter, K., Meiers, S., Kashin, S., Garrison, E., Auton, A., Lam, H. Y. K., Mu, X. J., Alkan, C., Antaki, D., Bae, T., Cerveira, E., Chines, P., Chong, Z., Clarke, L., Dal, E., Ding, L., Emery, S., Fan, X., Gujral, M., Kahveci, F., Kidd, J. M., Kong, Y., Lameijer, E. W., McCarthy, S., Flicek, P., Gibbs, R. A., Marth, G., Mason, C. E., Menelaou, A., Muzny, D. M., Nelson, B. J., Noor, A., Parrish, N. F., Pendleton, M., Quitadamo, A., Raeder, B., Schadt, E. E., Romanovitch, M., Schlattl, A., Sebra, R., Shabalin, A. A., Untergasser, A., Walker, J. A., Wang, M., Yu, F., Zhang, C., Zhang, J., Zheng-Bradley, X., Zhou, W., Zichner, T., Sebat, J., Batzer, M. A., McCarroll, S. A., Genomes Project, C., Mills, R. E., Gerstein, M. B., Bashir, A., Stegle, O., Devine, S. E., Lee, C., Eichler, E. E. & Korbel, J. O. 2015. An integrated map of structural variation in 2,504 human genomes. *Nature,* 526(7571)**,** pp 75-81.

Wakefield, J. 2009. Bayes factors for genome-wide association studies: comparison with P-values. *Genet Epidemiol,* 33(1)**,** pp 79-86.

Wang, K., Li, M. & Hakonarson, H. 2010. ANNOVAR: functional annotation of genetic variants from high-throughput sequencing data. *Nucleic Acids Res,* 38(16)**,** pp e164.

#### Supplementary Figures

**Figure S1. Quantile-Quantile (Q-Q) plot.** Observed versus expected -log10 *p*-values for the GWAS meta-analysis result (lambda=0.99).

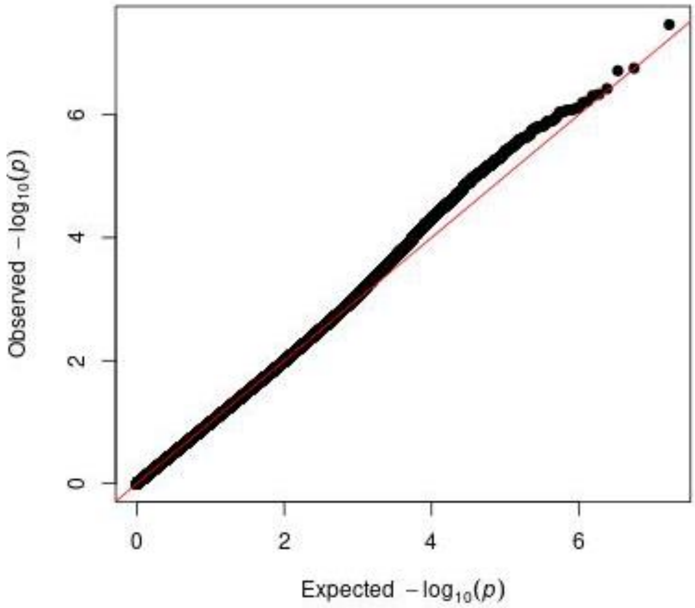

**Figure S2. Forest plots of the association analysis results for the nine prioritised variants.**

**rs749263563 (3p14.2)**

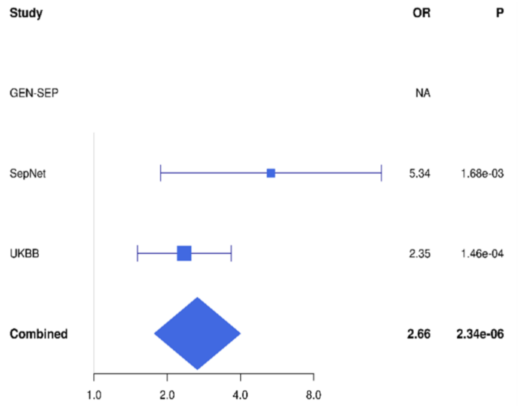


**rs183861060 (4q21.21)**

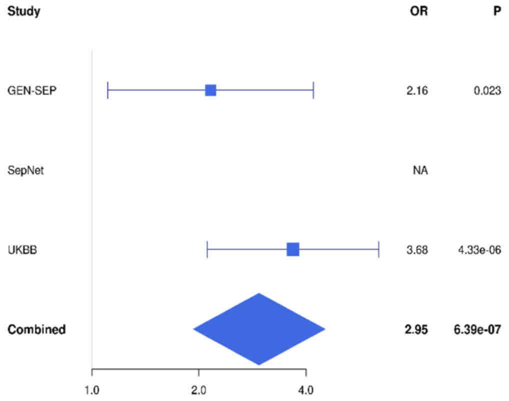


**rs116066418 (5q13.3)**

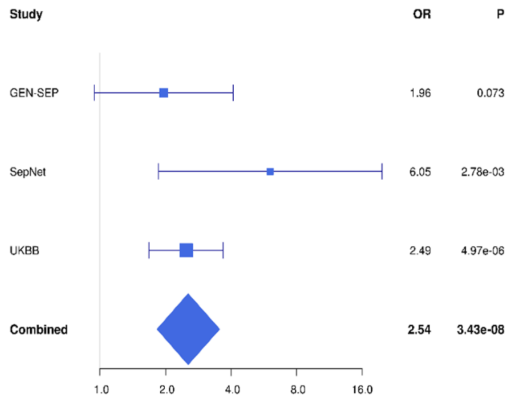


**rs59685452 (6q25.3)**

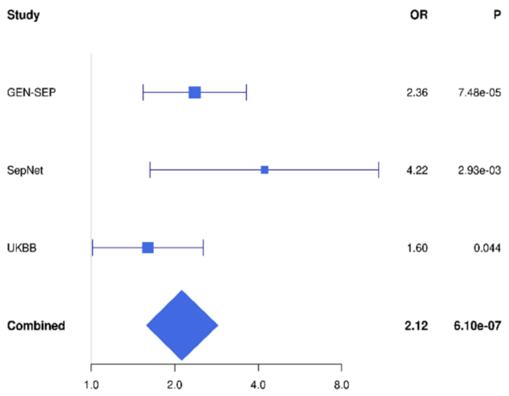


**rs12684841 (9q22.32)**

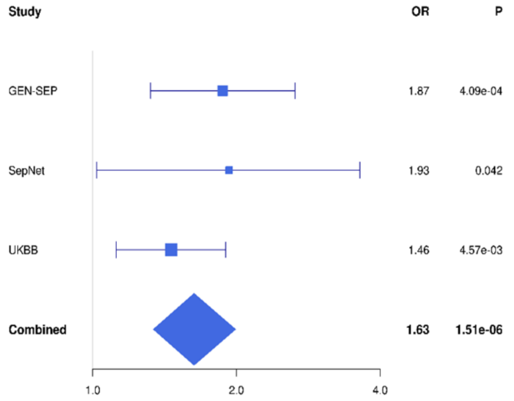


**rs11111647 (12q23.3)**

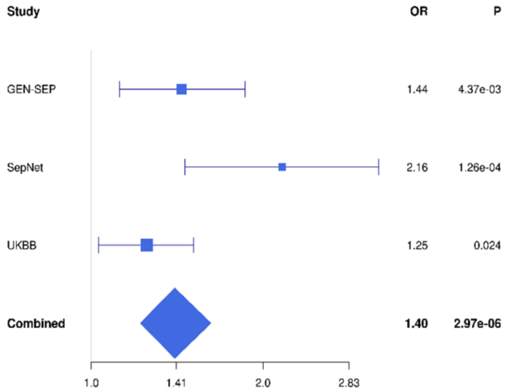

**rs118026254 (12q24.22)**

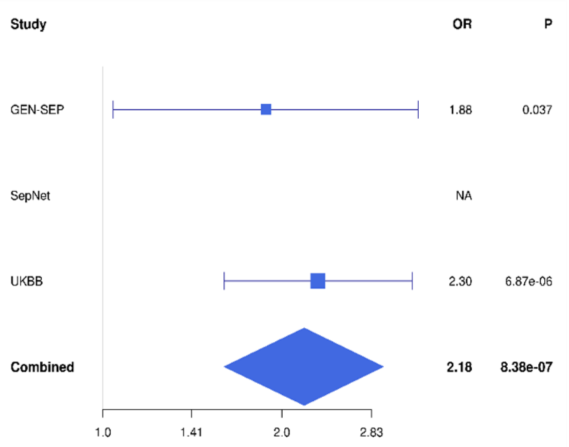


**rs8022645 (14q23.3)**

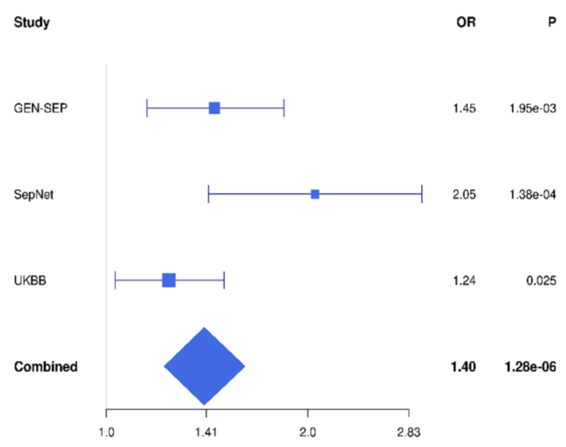


**rs4989808 (15q26.1)**

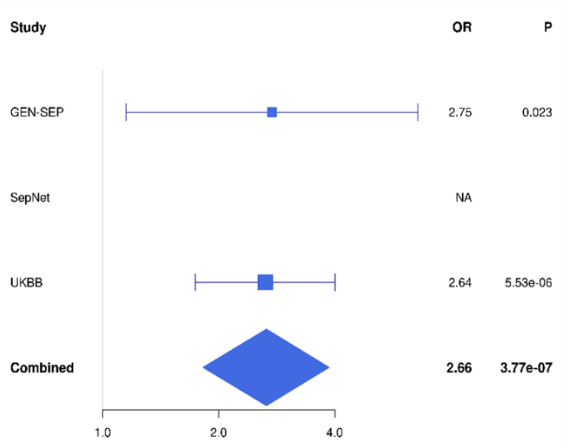

**Figure S3. Regional plots of the association results for the prioritised loci.** The y-axis shows the transformed *p*-values (-log10[*p*-value]), and the x-axis represents the chromosome positions (GRCh37/hg19). Genome-wide significance threshold (*p*-value=5.0x10$^{-8}$) is indicated by the horizontal dashed line. Linkage disequilibrium (LD) values ($r^2$) are based on the European population data from The 1000 Genomes Project and are represented according to the LD colour scheme of the top left legend. The plots were generated with LocusZoom (http://locuszoom.org/).

**rs749263563 (3p14.2)**

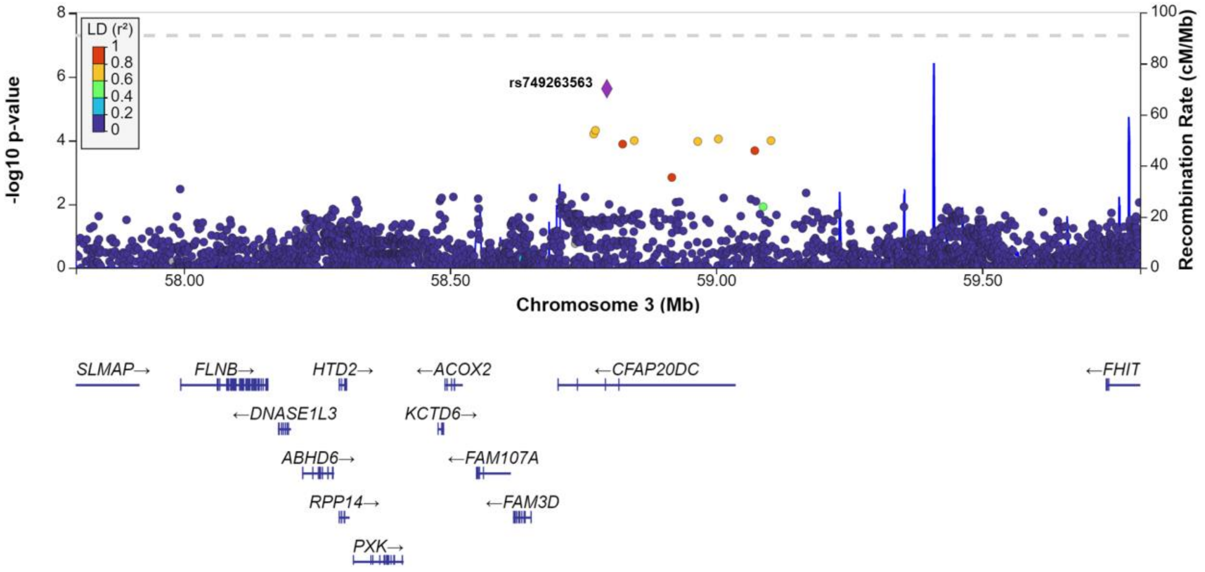


**rs183861060 (4q21.21)**

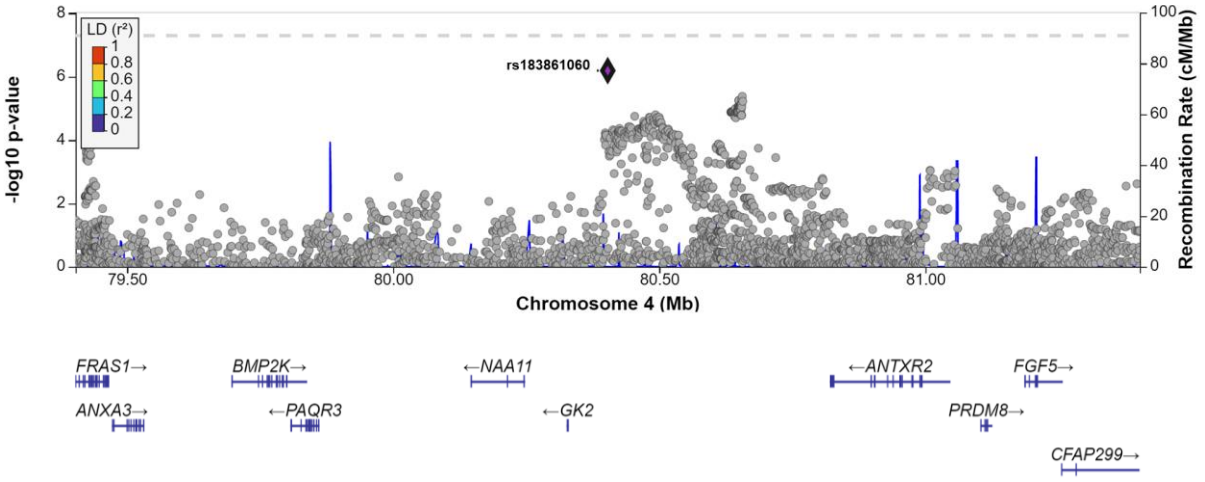

**rs116066418 (5q13.3)**

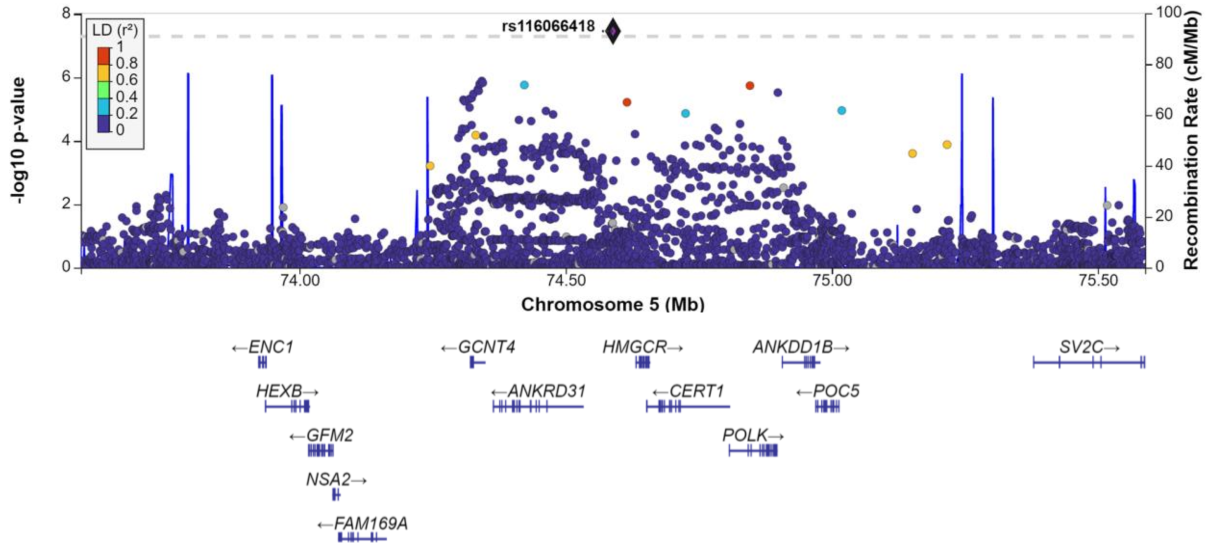


**rs59685452 (6q25.3)**

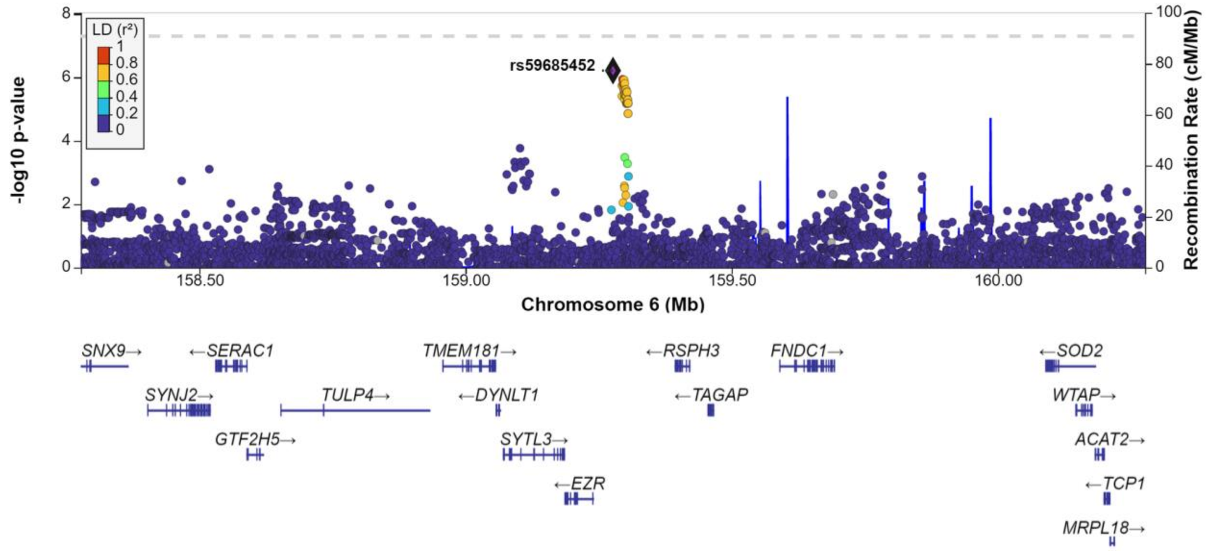


**rs12684841 (9q22.32)**

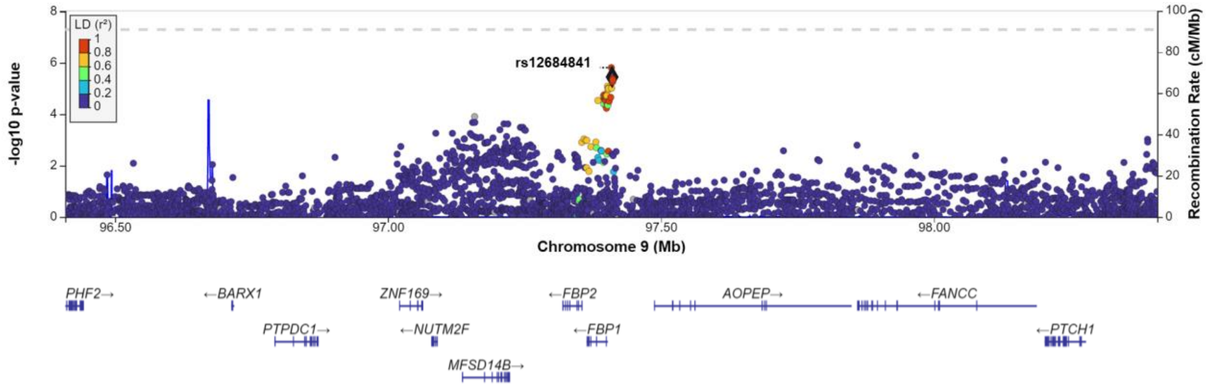

**rs11111647 (12q23.3)**

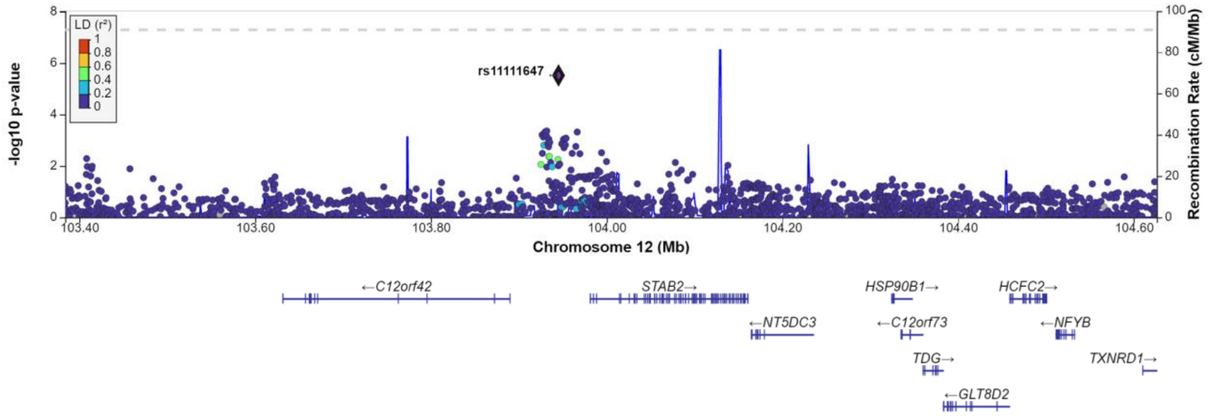


**rs118026254 (12q24.22)**

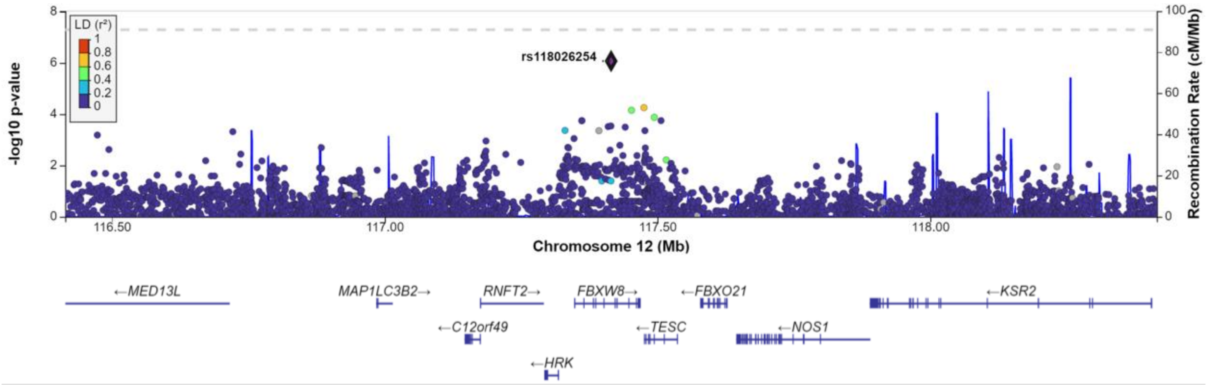


**rs8022645 (14q23.3)**

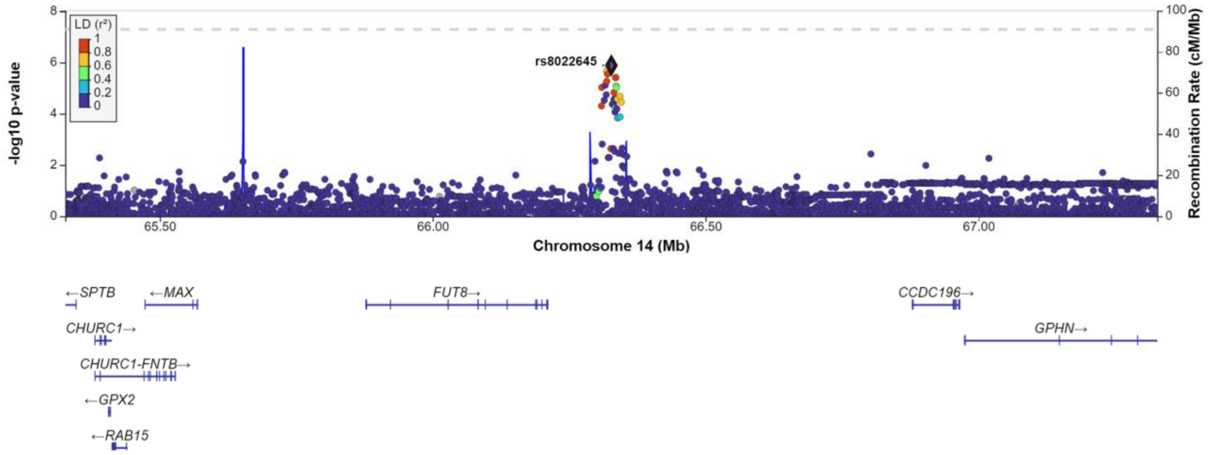

**rs4989808 (15q26.1)**

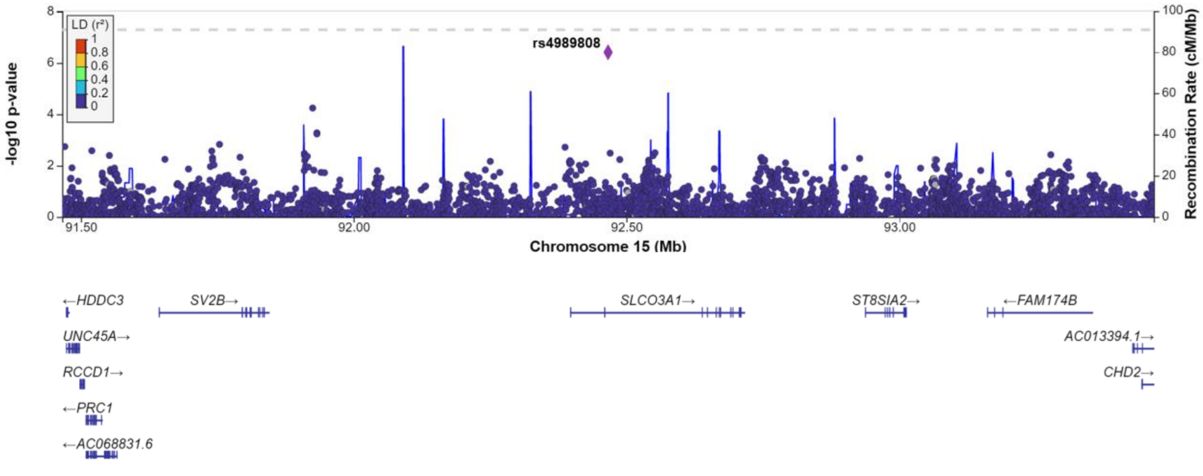

**Figure S4. Forest plot of the validation analysis results for rs9508032 (at *FLT1*) and rs7967111(at *BORCS5*).**

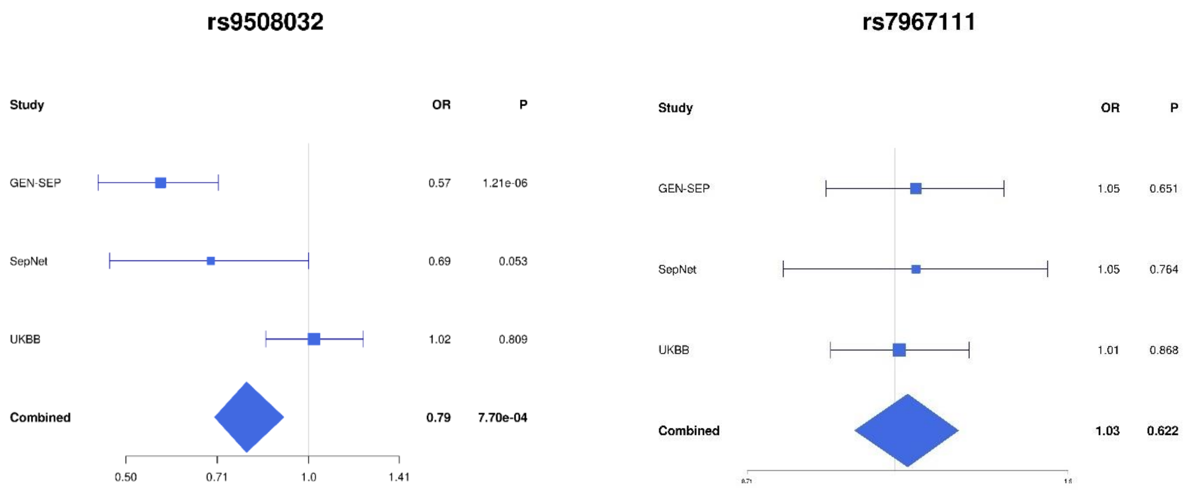

**Figure S5. Results of the Bayesian fine mapping for the prioritised loci.** The y-axis shows the transformed *p*-values ($-\log_{10}$ *p*-value), and the x-axis represents the chromosome positions (GRCh37/hg19). The orange diamond represents the sentinel variant of each region and the variants included in the credible sets are highlighted in green.

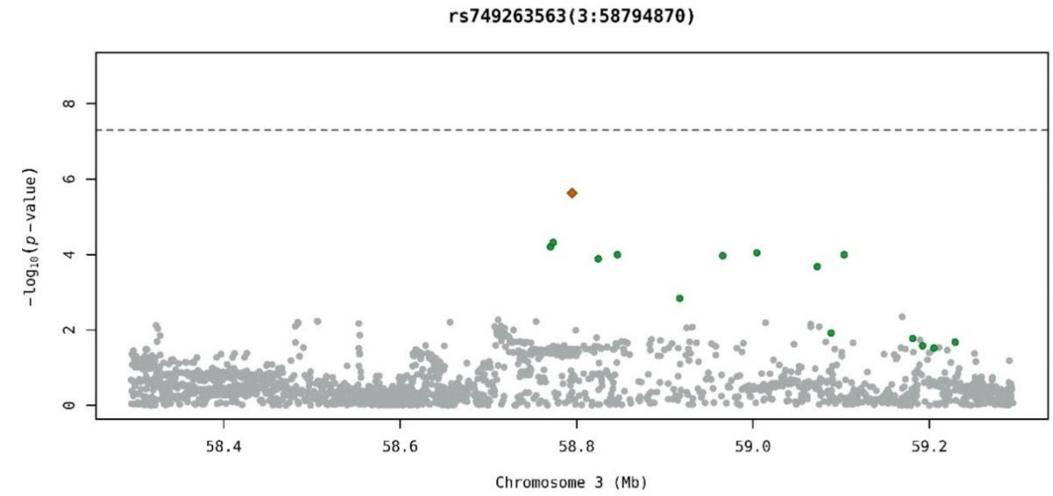


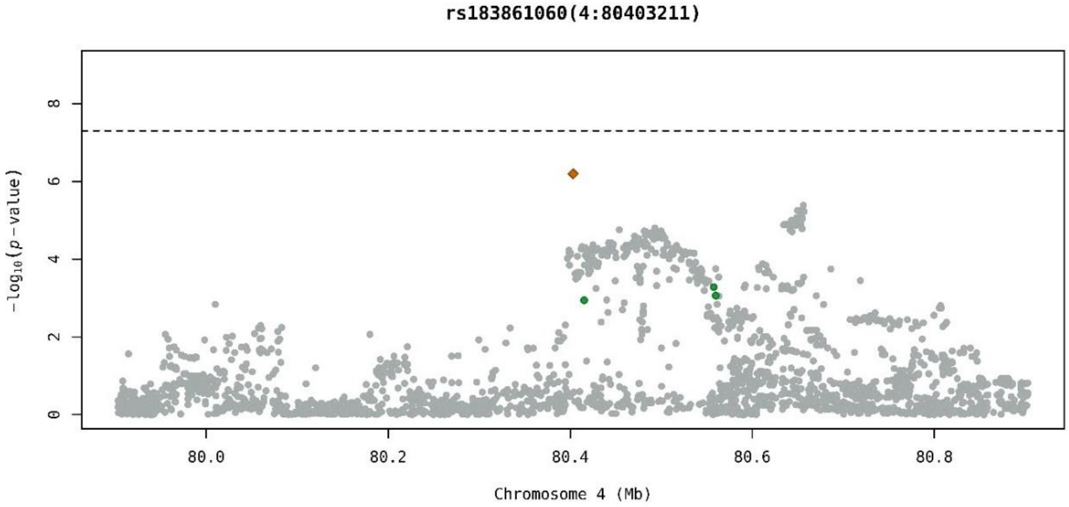


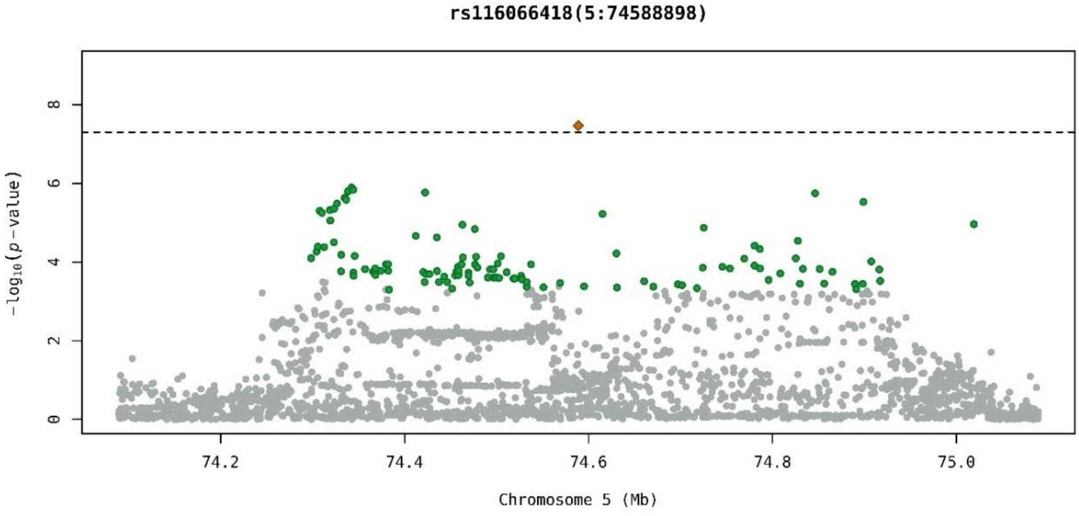

**rs59685452(6:159277046)**

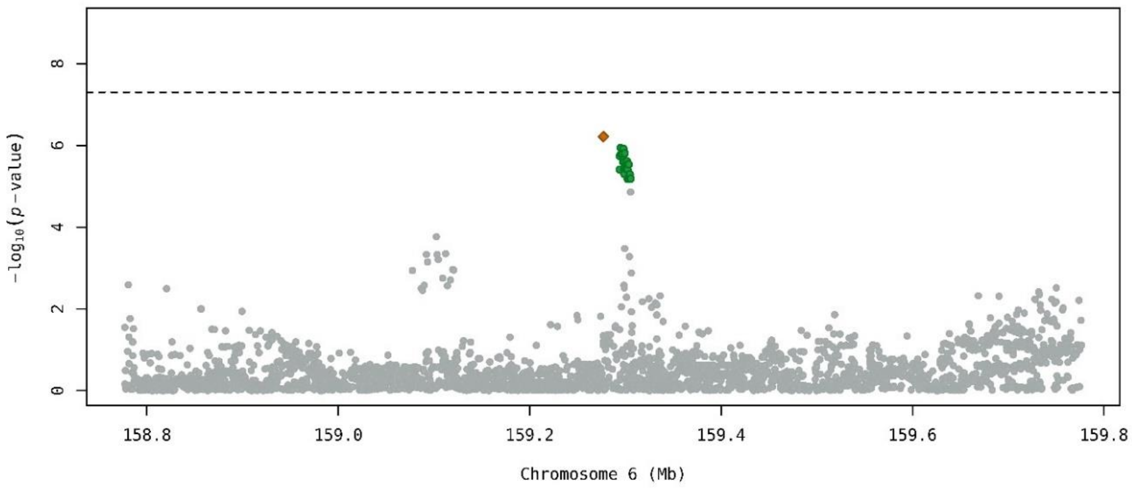


**rs12684841(9:97408971)**

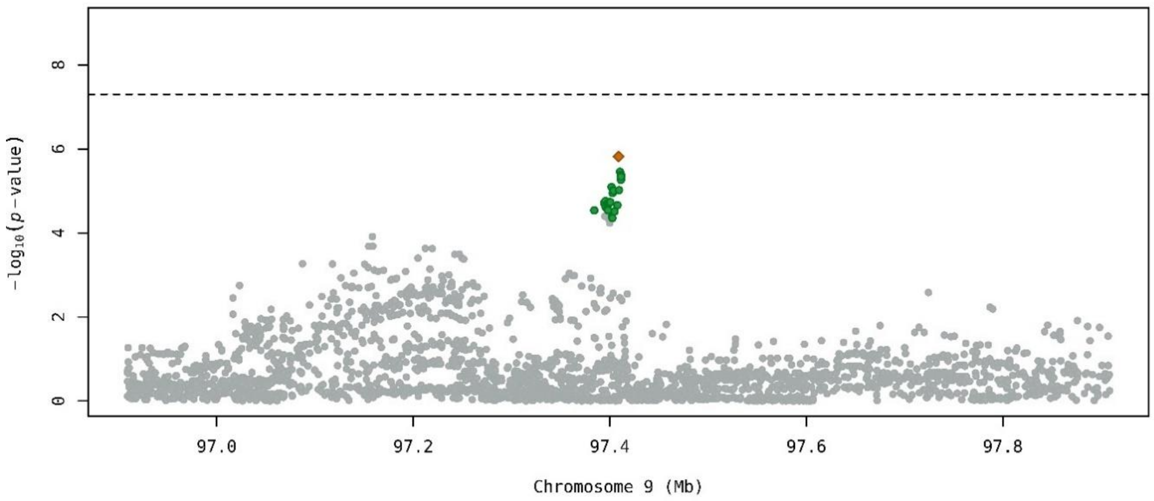


**rs11111647(12:103945224)**

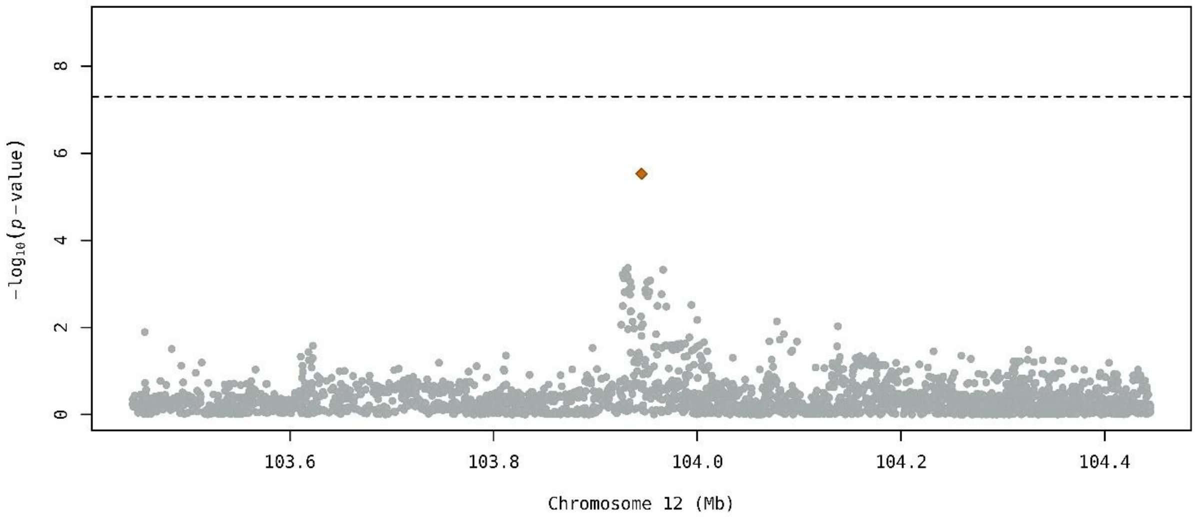

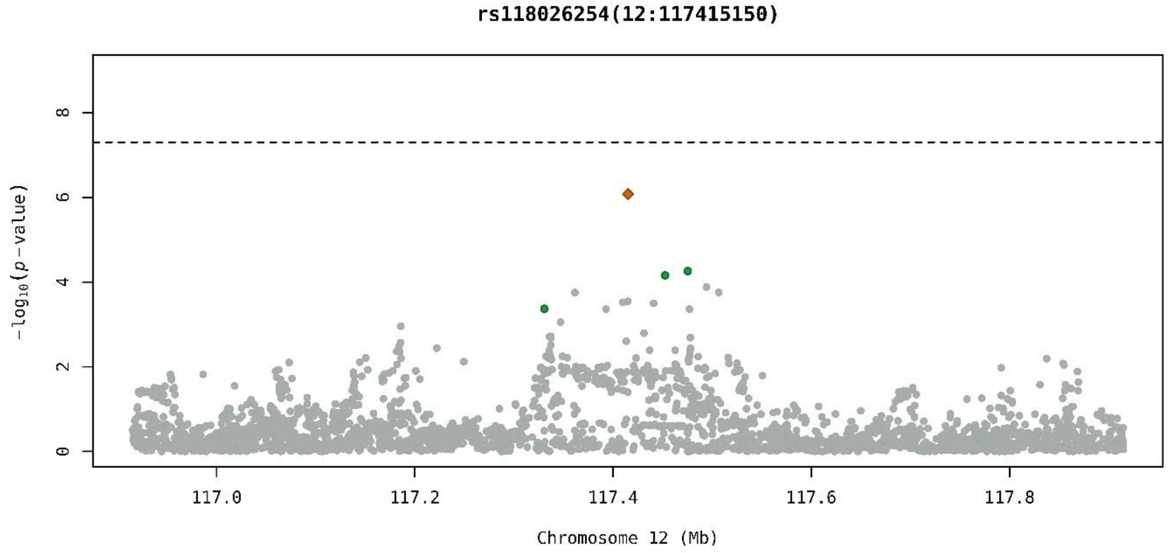
rs118026254(12:117415150)
−log10(p−value)
0
2
4
6
8
117.0
117.2
117.4
117.6
117.8
Chromosome 12 (Mb)

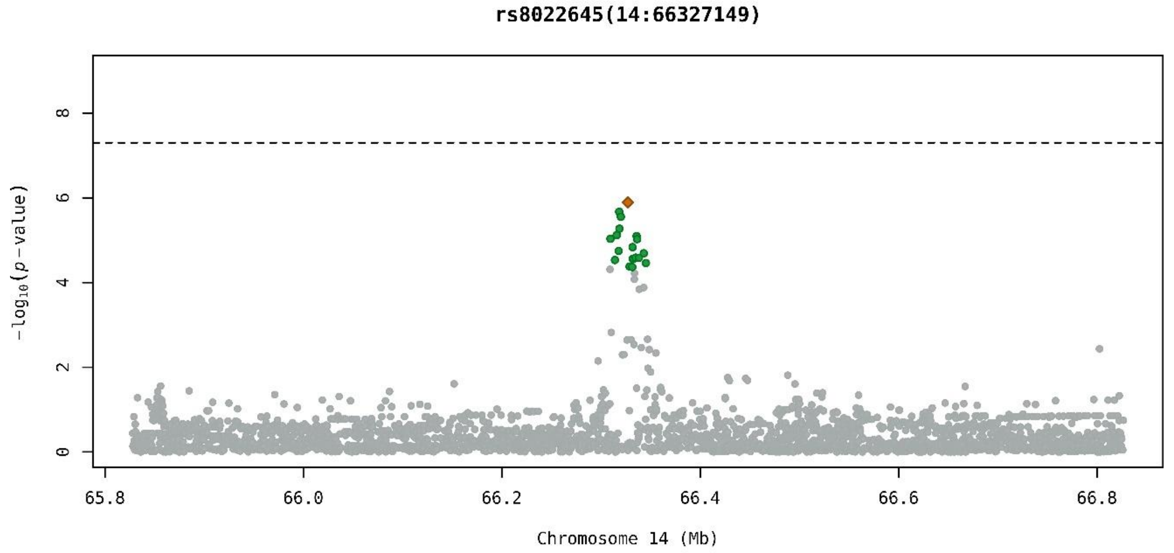
rs8022645(14:66327149)
−log10(p−value)
0
2
4
6
8
65.8
66.0
66.2
66.4
66.6
66.8
Chromosome 14 (Mb)

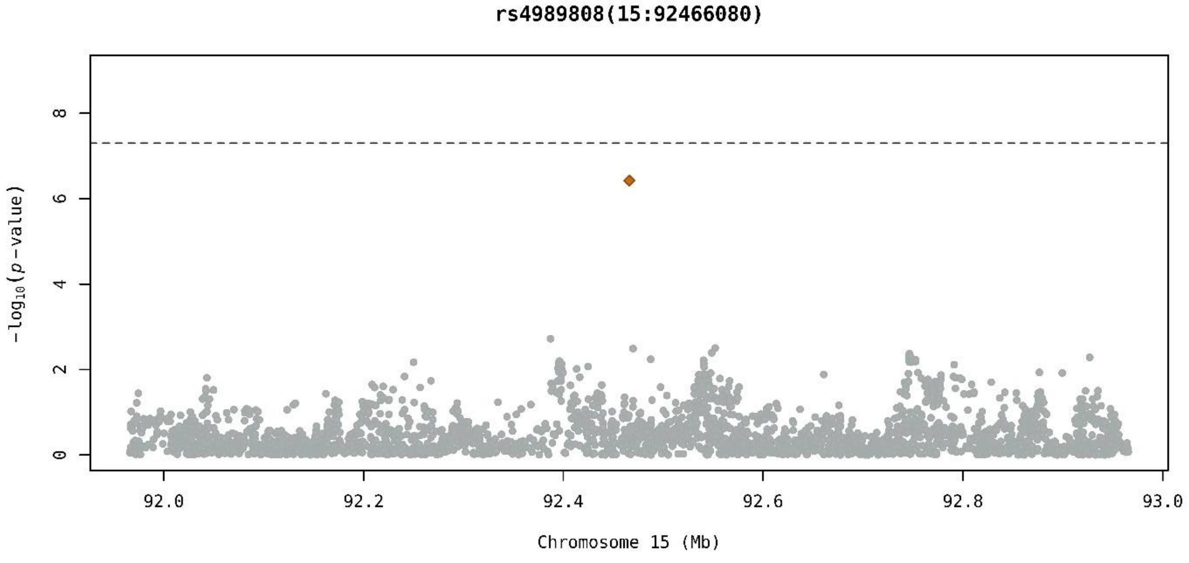
rs4989808(15:92466080)
−log10(p−value)
0
2
4
6
8
92.0
92.2
92.4
92.6
92.8
93.0
Chromosome 15 (Mb)

#### Supplementary Tables

##### Table S1. ICD-10 codes used in the UK Biobank study.

| ARDS cause | ICD-10 Codes/ Data-Field 41270 | Definition |
|---|---|---|
| Sepsis | A02.1 | *Salmonella* septicaemia |
| | A20.7 | Septicemic plague |
| | A22.7 | Anthrax septicaemia |
| | A32.7 | Listerial septicaemia |
| | A40 | Streptococcal septicaemia |
| | A40.0 | Septicaemia due to *Streptococcus*, group A |
| | A40.1 | Septicaemia due to *Streptococcus*, group B |
| | A40.2 | Septicaemia due to *Streptococcus*, group D |
| | A40.3 | Septicaemia due to *Streptococcus pneumoniae* |
| | A40.8 | Other streptococcal septicaemia |
| | A40.9 | Streptococcal septicaemia, unspecified |
| | A41 | Other septicaemia |
| | A41.0 | Septicaemia due to *Staphylococcus aureus* |
| | A41.1 | Septicaemia due to other specified *Staphylococcus* |
| | A41.2 | Septicaemia due to unspecified *Staphylococcus* |
| | A41.3 | Septicaemia due to *Haemophilus influenzae* |
| | A41.4 | Septicaemia due to anaerobes |
| | A41.5 | Septicaemia due to other Gram-negative organisms |
| | A41.8 | Other specified septicaemia |
| | A41.9 | Septicaemia, unspecified |
| | A42.7 | Actinomycotic septicaemia |
| | B37.7 | Candidal septicaemia |
| | R57.2 | Septic shock |
| | R65.1 | Systemic Inflammatory Response Syndrome of infectious origin with organ failure |
| Infections due to complications of surgical and medical care | T80.2 | Infections following infusion, transfusion and therapeutic injection |
| | T81.4 | Infection following a procedure, not elsewhere classified |
| | T82.6 | Infection and inflammatory reaction due to cardiac valve prosthesis |
| | T82.7 | Infection and inflammatory reaction due to other cardiac and vascular devices, implants and grafts |
| | T83.5 | Infection and inflammatory reaction due to prosthetic device, implant and graft in urinary system |

| | | |
|---|---|---|
| | T83.6 | Infection and inflammatory reaction due to prosthetic device, implant and graft in genital tract |
| | T84.7 | Infection and inflammatory reaction due to other internal orthopaedic prosthetic devices, implants and grafts |
| | T85.7 | Infection and inflammatory reaction due to other internal prosthetic devices, implants and grafts |
| | T88.0 | Infection following immunisation |
| Pneumonia | J10.0 | Influenza with pneumonia, influenza virus identified |
| | J11.0 | Influenza with pneumonia, virus not identified |
| | J12 | Viral pneumonia, not elsewhere classified |
| | J13 | Pneumonia due to *Streptococcus pneumoniae* |
| | J14 | Pneumonia due to *Haemophilus influenzae* |
| | J15 | Bacterial pneumonia, not elsewhere classified |
| | J16 | Pneumonia due to other infectious organisms, not elsewhere classified |
| | J17 | Pneumonia in diseases classified elsewhere |
| | J18 | Pneumonia, organism unspecified |
| | J22 | Unspecified acute lower respiratory infection |
| Pancreatitis | K85 | Acute pancreatitis |
| Patients with ARDS associated to head, chest or other major injury (= Severe trauma) | T79 | Certain early complications of trauma |
| | T00-T07 | Injuries involving multiple body regions |
| | S02 | Fracture of skull and facial bones |
| | S04 | Injury of cranial nerves |
| | S06 | Intracranial injury |
| | S07 | Crushing injury of head |
| | S08 | Traumatic amputation of part of head |
| | S097 | Multiple injuries of head |
| | S22 | Fracture of rib(s), sternum and thoracic spine |
| | S23.0 | Traumatic rupture of thoracic intervertebral disc |
| | S24 | Injury of nerves and spinal cord at thorax level |
| | S28 | Crushing injury of thorax and traumatic amputation of part of thorax |
| | S29.7 | Multiple injuries of thorax |
| Inhalation of harmful substances | W78 | Inhalation of gastric contents |
| | T58 | Toxic effect of carbon monoxide |
| | T59 | Toxic effect of other gases, fumes and vapours |
| Poisoning | T36-T50 | Poisoning by drugs, medicaments and biological substances |
| | X61 | Intentional self-poisoning by and exposure to antiepileptic, sedative-hypnotic, antiparkinsonism and psychotropic drugs, not elsewhere classified |

**Table S2. Genetic ancestry of individuals in the UK Biobank study.**

| Ancestry | % |
|---|---|
| African | 1.25 |
| Asian | 3.12 |
| Chinese | 0.62 |
| European | 92.52 |
| Mixed (black & white) | 0.62 |
| Mixed (others) | 1.87 |

**Table S3. Bayesian fine mapping and SNP deleteriousness assessment**

| Locus | Credible set | Highest PP from the credible set | Sentinel variant | | | Highest phenotypic impact variant* | | | |
|---|---|---|---|---|---|---|---|---|---|
| | | | rsid | PP | CADD score | rsid | $r^2$ | PP | CADD score |
| **3p14.2** | 15 | 0.218 | rs749263563 | 0.218 | 0.39 | rs80308704 | 0.89 | 0.081 | 17.80 |
| **4q21.21** | 4 | 0.735 | rs183861060 | 0.735 | 6.72 | rs183861060 | 1.00 | 0.735 | 6.72 |
| **5q13.3** | 119 | 0.090 | rs116066418 | 0.060 | 3.18 | rs6893216 | 0.18 | 0.002 | 16.66 |
| **6q25.3** | 33 | 0.079 | rs59685452 | 0.079 | 0.87 | rs75891667 | 0.92 | 0.032 | 14.33 |
| **9q22.32** | 28 | 0.150 | rs12684841 | 0.150 | 1.46 | rs73523269 | 0.96 | 0.056 | 15.78 |
| **12q23.3** | 1 | 0.972 | rs11111647 | 0.972 | 5.44 | rs11111647 | 1.00 | 0.972 | 5.44 |
| **12q24.22** | 4 | 0.795 | rs118026254 | 0.795 | 0.31 | rs117510704 | 0.41 | 0.033 | 5.67 |
| **14q23.3** | 19 | 0.231 | rs8022645 | 0.231 | 2.15 | rs12896836 | 0.39 | 0.018 | 13.71 |
| **15q26.1** | NA | NA | rs4989808 | NA | 7.77 | rs4989808 | 1.00 | NA | 7.77 |

CADD: combined annotation dependent depletion v.1.6; PP: Posterior probabilities; $r^2$: linkage disequilibrium between the sentinel variant and the SNP with the highest phenotypic impact in the locus. *Prioritised SNP based on the top-ranked CADD score for each locus.

**Table S4. Highest phenotypic impact SNPs follow-up: first-stage meta-analysis association results, gene expression and PheWAS**

| Sentinel variant [locus] | Highest phenotypic impact SNP* | chr:position (GRCh37/hg19) | NEA/EA | OR (95%CI) | *p*-value | Nearest gene/s | eQTLs# | PheWAS (*p*<0.005) |
|---|---|---|---|---|---|---|---|---|
| rs749263563, [**3p14.2**] | rs80308704 | 3:58965636 | G/T | 1.88 (1.37, 2.59) | $1.07x10^{-4}$ | *CFAP20DC* | NA | Vascular diseases of the intestine, Disorders involving the immune mechanism, Arrhythmia, Systemic sclerosis, Human herpes virus 6 p101k antibody levels, Lung volume |
| rs183861060, [**4q21.21**] | rs183861060 | 4:80403211 | A/G | 2.95 (1.93, 4.52) | $6.39x10^{-7}$ | *GK2* | NA | Sialic acid-binding Ig-like lectin 7 levels, Chronic liver disease and cirrhosis, Acute pancreatitis, Diseases of pancreas |
| rs116066418, [**5q13.3**] | rs6893216 | 5:74442964 | T/C | 1.38 (1.16, 1.65) | $2.35x10^{-4}$ | *ANKRD31* | + *POC5* (Cultured fibroblasts, Thyroid, EBV-transformed lymphocytes). H4 = <5%; H3 = 87%<br><br>+ *ANKDD1B* (Artery-Tibial). H4 = 71.50% [colocalise]; H3 = 25.20%<br><br>+ *ANKDD1B* (Artery-Aorta). H4 = 28.20%; H3= 60.30% | Low density lipoprotein cholesterol levels, Total cholesterol levels, Apolipoprotein B levels, Statin medication, Platelet count, Mean platelet volume, C-reactive protein levels, Disorders of lipoid metabolism, Diastolic blood pressure, Monocyte percentage of white cells, Cardiovascular disease, Immature fraction of reticulocytes, Neutrophil count, Non-allergic asthma |
| rs59685452, [**6q25.3**] | rs75891667 | 6:159301294 | A/G | 2.05 (1.52, 2.77) | $2.43x10^{-6}$ | *EZR* | NA | Other and unspecified coagulation defects, Hypotension, Interleukin-18 levels, Symptoms and signs involving the circulatory and respiratory systems, Other bacterial diseases, Bacterial infection other or unspecified, Other specific/unspecified arthritis, Neutrophil count |
| rs12684841, [**9q22.32**] | rs73523269 | 9:97411359 | T/C | 1.59 (1.30, 1.94) | $5.44x10^{-6}$ | *FBP1, AOPEP* | - *FBP1* (Oesophagus-Mucosa). H4 = 88.80% [colocalise]; H3 = 5.41%<br><br>+ *MFSD14B* (Whole Blood) H4 = 5.36%; H3 = 74.30% | Epstein-Barr virus VCA p18 antibody levels, Triglyceride levels, Viral hepatitis b, Red blood cell count, Pulmonary oedema, Chronic hepatitis, Chronic rhinitis nasopharyngitis and pharyngitis, Cardiac problem, Red blood cell count, Interleukin-6 receptor subunit alpha levels |
| rs11111647, [**12q23.3**] | rs11111647 | 12:103945224 | G/A | 1.40 (1.22, 1.62) | $2.97x10^{-6}$ | *STAB2* | NA | Alloprevotella abundance, Platelet endothelial cell adhesion molecule levels, infectious diseases, Rheumatism and fibrositis |

| Sentinel variant [locus] | Highest phenotypic impact SNP* | chr:position (GRCh37/hg19) | NEA/EA | OR (95%CI) | *p*-value | Nearest gene/s | eQTLs# | PheWAS (*p*<0.005) |
|---|---|---|---|---|---|---|---|---|
| rs118026254 [**12q24.22**] | rs117510704 | 12:117330700 | T/C | 1.56 (1.22, 1.99) | $4.27 \times 10^{-4}$ | *HRK* / *FBXW8* | NA | Pulmonary heart disease, diseases of pulmonary circulation, Childhood asthma (age<16), Pulmonary embolism, Pulmonary heart disease, Bronchopneumonia and lung abscess, Mean platelet (thrombocyte) volume, Allergic rhinitis, Asthma and allergy, Pleural plaque |
| rs8022645 [**14q23.3**] | rs12896836 | 14:66332007 | C/T | 1.29 (1.15, 1.45) | $2.74 \times 10^{-5}$ | *FUT8* / *CCDC196* | NA | Fatty acid levels, High cholesterol, Type 2 diabetes, Interleukin-1 receptor type 1 measurement, Recent medication for bronchiectasis, Other acute lower respiratory infections |
| rs4989808 [**15q26.1**] | rs4989808 | 15:92466080 | A/G | 2.66 (1.82, 3.87) | $3.77 \times 10^{-7}$ | *SLCO3A1* | NA | Lymphocyte and Neutrophil percentage of white cells, Abnormal sputum, Bronchopneumonia and lung abscess, VIH |

*Prioritised SNP based on the top-ranked CADD score for each locus. # Increased (+) or decreased (-) gene expression. Directions of effect refer to the ARDS risk allele. The H4 values inform the posterior probability of colocalisation between the ARDS risk signal and the gene expression eQTL signal in the tissues stated. A H4>70% indicates that both traits are associated and share a single causal variant, while A H3>70% indicates that both traits are associated, but with different causal variants. CADD: combined annotation dependent depletion v.1.6, chr: chromosome, CI: confidence interval, EBP: Epstein-Barr Virus; eQTLs: expression quantitative locus, based on Genotype-Tissue Expression (GTEx) project V8 (https://www.gtexportal.org/home/); NEA/EA: non-effect allele/effect allele; OR: odds ratio, PheWAS: Phenome-wide association studies based on reported association p-value<0.005 for the prioritised SNPs (https://genetics.opentargets.org/).
